# Unlocking low- and middle-income countries to detect SARS-CoV-2

**DOI:** 10.1101/2021.04.14.21255479

**Authors:** Roberto Alcántara, Katherin Peñaranda, Gabriel Mendoza, Jose A. Nakamoto, Johanna Martins-Luna, Juana del Valle, Vanessa Adaui, Pohl Milón

**Affiliations:** Centre for Research and Innovation, Faculty of Health Sciences, Universidad Peruana de Ciencias Aplicadas (UPC), Lima, Peru; Laboratorio de Biología Molecular, Instituto de Investigación Nutricional, Lima, Peru

**Keywords:** Covid-19, SARS-CoV-2, LMICs, CRISPR, Fluorescence, Molecular Testing

## Abstract

Low- and middle-income countries (LMICs) are significantly affected by SARS-CoV-2, partially due to their limited capacity for local development of molecular testing and accentuated by the international supply shortage. Here, we describe a molecular toolkit that can be readily produced and deployed in LMICs using minimal laboratory equipment. Our results show that mid-scale production of enzymes and nucleic acids can supply thousand tests per production batch. One-step RT-PCR was optimized for two SARS-CoV-2 loci and coupled to CRISPR/Cas12a detection. The clinical validation indicated a sensitivity near 100% for mid and high viral load samples (Cq ≤ 33). The specificity was around 100% regardless of viral load. The toolkit was used with the mobile laboratory BentoLab, potentially unlocking LMICs to implement detection services in unattended regions. Altogether, we provide detailed methods and performance evidence of molecular tools aiming to aid LMICs to deploy molecular testing for current or future pathogenic outbreaks.

**One Sentence Summary:** We describe a molecular toolkit for the detection of SARS-CoV-2 that is compatible with available facilities in low- and middle-income countries (LMICs).

## Introduction

SARS-CoV-2 has infected more than 113 million people and caused 2,526,007 deaths worldwide, forcing countries to adopt strict quarantine measures to control the contagion (as of March 2021, WHO 2021). This has negatively impacted local and global economies, with more than trillions of dollars in economic loss (Ibn-Mohammed et al., 2021). Since the beginning of the Covid-19 pandemics, early detection of the virus appeared evident in order to restrict the rate of transmission, extremely high for SARS-CoV-2. High-income countries (HICs) implemented massive molecular testing facilities together with contact tracing technologies and proper isolation of positive patients (Ferretti et al., 2020; Kucharski et al., 2020). Consequently, the scale of the pandemic outbreak generated an unprecedented high demand for molecular testing equipment and materials, precluding their access in low- and middle-income countries (LMICs) (Figure 1A). The shortage of viral RNA extraction kits, quantitative reverse transcription polymerase chain reaction (RT-qPCR) materials and instrumentation limited the capability of LMICs to accurately identify carriers of the virus (Adepoju, 2020; Rubin et al., 2020). Both the lack of molecular testing and the high rate of viral propagation configured into a perfect storm for LMICs(Abbott et al., 2020). Countries such as Brazil, Argentina, Colombia, Peru, among others, surpassed developed countries in both the number of infections and deaths per inhabitant (Roser and Ortiz-Ospina 2021). Consequently, there is a high need to habilitate LMICs with tools for SARS-CoV-2 detection, aiming to decrease their dependence on the international supply.

**Figure 1:**
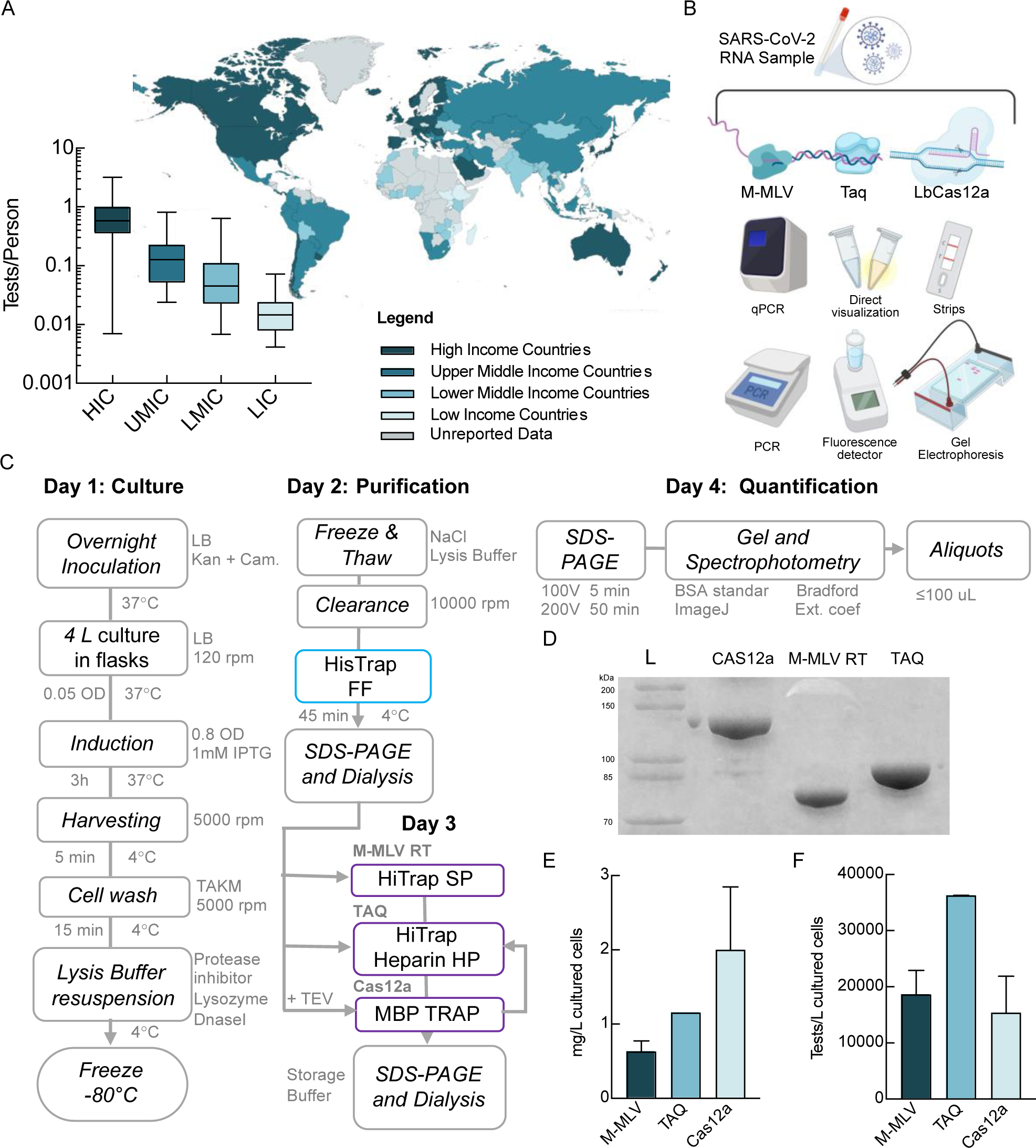
SARS-CoV-2 testing availability, inequality, and local production of reagents. (A) Molecular testing availability as function of income classification by the World Bank of countries (Roser and Ortiz-Ospina 2021). (B) Current molecular diagnostic platforms for detection of SARS-CoV-2. (C) Production scheme for recombinant DNA polymerases Taq and M-MLV and LbCas12a. (D) Recombinant enzyme visualization and purity evaluation by SDS-PAGE 10%. (E) Comparison of enzyme yield expressed as mg of pure protein produced in 1 L of bacterial cell culture. (F) Estimated total test (reverse transcription, PCR or CRISPR/Cas12a) reactions achievable in 1 L of bacterial cell culture producing recombinant enzymes M-MLV reverse transcriptase (RT), Taq DNA polymerase or LbCas12a, respectively, out of *E. coli.* Error bars in (E) and (F) show the standard error while in (A) indicate the minimum and maximum.

In addition to the high demand for biotechnological supplies, LMICs have limited infrastructure, laboratory equipment, and trained professionals. This is further evidenced in geographical regions that are distant from major capital cities. Several new methods have been recently reported, using RT-qPCR (Corman et al., 2020), LAMP (Hu et al., 2020), CRISPR/Cas (Hou et al., 2020), among others (Figure 1B). Despite their potential for coping with LMICs’ limitations is high, a concrete solution is yet unavailable. RT-qPCR requires real-time thermocyclers that are costly and not readily available. LAMP-based methods require basic laboratory equipment, yet their deployment in LMICs use commercially available enzymes or kits rather than local production. CRISPR-based methods are still due to be widely used worldwide, albeit several proofs of concept are available (Broughton et al., 2020; Hou et al., 2020; Metsky et al., 2020). Available CRISPR-based methods can lay into two categories, detection after amplification of the genetic material and direct detection by the CRISPR/Cas complex (Fozouni et al., 2021). Detection after amplification of the genetic material can be also divided into two categories depending on the amplification method: isothermal (LAMP or RPA) (Hu et al., 2020; Joung et al., 2020; Sun et al., 2021; Wang et al., 2020) or by thermal cycling (reverse transcription plus PCR) (Huang et al., 2020). Thus, CRISPR/Cas provides a versatile tool for detecting the amplified genetic material from SARS-CoV-2.

Here, we describe a molecular toolkit that can be readily produced and deployed in LMICs using minimal and broadly available laboratory equipment (Figure 1B). Together with detailed protocols for the production of biologicals and step by step optimization (Figure 1C), we provide a clinical performance evaluation for SARS-CoV-2 detection. Altogether, we sought to alleviate the reduced availability of molecular detection methods that affect LMICs in the control of the Covid-19 pandemic (Figure 1A).

## Results

### Local production of key biological components

The current pandemic situation caused by SARS-CoV-2 has revealed the vast number of limitations of LMICs health systems. In response to this need, we first validated and adapted open-source protocols for the local production of biological reagents using minimal laboratory equipment in LMICs focusing on versatile and widely used enzymes, namely the Taq DNA polymerase and M-MLV reverse transcriptase (M-MLV RT), for nucleic acid amplification (Figure 1C). Some of the methods in our toolkit use the LbCas12a (from herein Cas12a) enzyme for CRISPR-Cas-mediated detection. Thus, we also describe the production of Cas12a together with the required CRISPR guide RNAs (crRNA) in a LMIC setup (Figure S1).

Each required enzyme was produced to high purity following a four-day scheme (Figure 1C). On day one, *E. coli* BL21 (DE3) carrying the respective expression plasmid (Key Resources Table) was grown in Luria-Bertani medium (typically 1-4 L) using antibiotic selection throughout the process. Protein induction was achieved by the addition of IPTG. Typically, 3-5 g of dry cells were obtained for each liter of cell culture. Days two and three involved purification steps, which can be carried out manually or in a scaling-up stage using liquid chromatography equipment (FPLC or HPLC) if available. Typically, 6-12 mL of enzymes were obtained and stored. The final day is dedicated to enzyme quantification and characterization through gel imaging and spectrophotometry, preparation of aliquots and storage of the produced enzymes at low temperatures (-20°C to -80 °C) (Figure 1C, Detailed protocols are available in Star Methods). The standardized protocols showed high purity and yields of the enzymes (Figures 1D-F). Average purity ranged between 90% and 99%, comparable to or higher than commercial counterparts (Figure 1D). The protein production yield unlocks between 20 and 40 thousand RT-PCR reactions per liter of cell culture (Figure 1F). The production of Cas12a allows around 20 thousand CRISPR-Cas reactions per liter of cell culture. As seen in Figure 1 E, Cas12a presents the highest yield with 2 mg/L cell, followed by Taq DNA polymerase with 1.2 mg/L cell and finally, M-MLV RT with 0.6 mg/L cell. However, due to the difference in working concentrations, Taq allows the highest number of reactions per liter of cell culture.

In general, upscaling the production process to 4 L cell culture allows enzyme production for 160 thousand PCR reactions, 80 thousand retro-transcription reactions and 60 thousand CRISPR-Cas12a reactions. crRNAs can also be produced locally from synthetic dsDNA templates (File S1) using commercially available *in vitro* transcription kits (Key Resources Table). crRNA production and purification from a 40 µL reaction yielded on average 340 µg of *in vitro* transcribed crRNA, accounting for more than 16 thousand CRISPR/Cas12a-mediated detection reactions. Altogether, the methods described in detail are compatible with the timely production of all key components for molecular detection of SARS-CoV-2 in a minimal laboratory setup (Star Methods).

### Target selection and primer design

Currently, nucleic acid amplification tests (NAAT) are considered the recommended, most sensitive tests for SARS-CoV-2 detection (Böger et al., 2021; Kevadiya et al., 2021). RT-qPCR is the gold standard molecular test (Corman et al., 2020) and different targets at the ORF1ab, S, E, or N gene have been reported for molecular detection (Broughton et al., 2020; Esbin et al., 2020; Huang et al., 2020; Javalkote et al., 2020). Differences in specificity, amplification efficiency, and also in downstream detection (i.e. CRISPR/Cas) have been described (Joung et al., 2020; Xiong et al., 2020). Here, we selected one target at each, 5’ and 3’, end of the SARS-CoV-2 genome, aiming to detect infectious viral RNA rather than subgenomic RNA or partially degraded molecules (Kim et al., 2020) (Figure 2A). Multiple alignment analysis of available viral genomes highlighted regions in the ORF1ab and N genes that were highly conserved (File S1). Additionally, the potential detection *loci* encountered were screened for structured RNA segments and CRISPR/Cas12a compatible sequences (File S2). The conserved region II (CII) located within the ORF1ab gene complied with the above premises and was used in this study. Additionally, a previously reported sequence was selected for the N gene (N) (Broughton et al., 2020). These targets were compared against the reported genome variants of SARS-CoV-2 and no mutations have been observed yet (Koyama et al., 2020) (Figure 2A).

**Figure 2.**
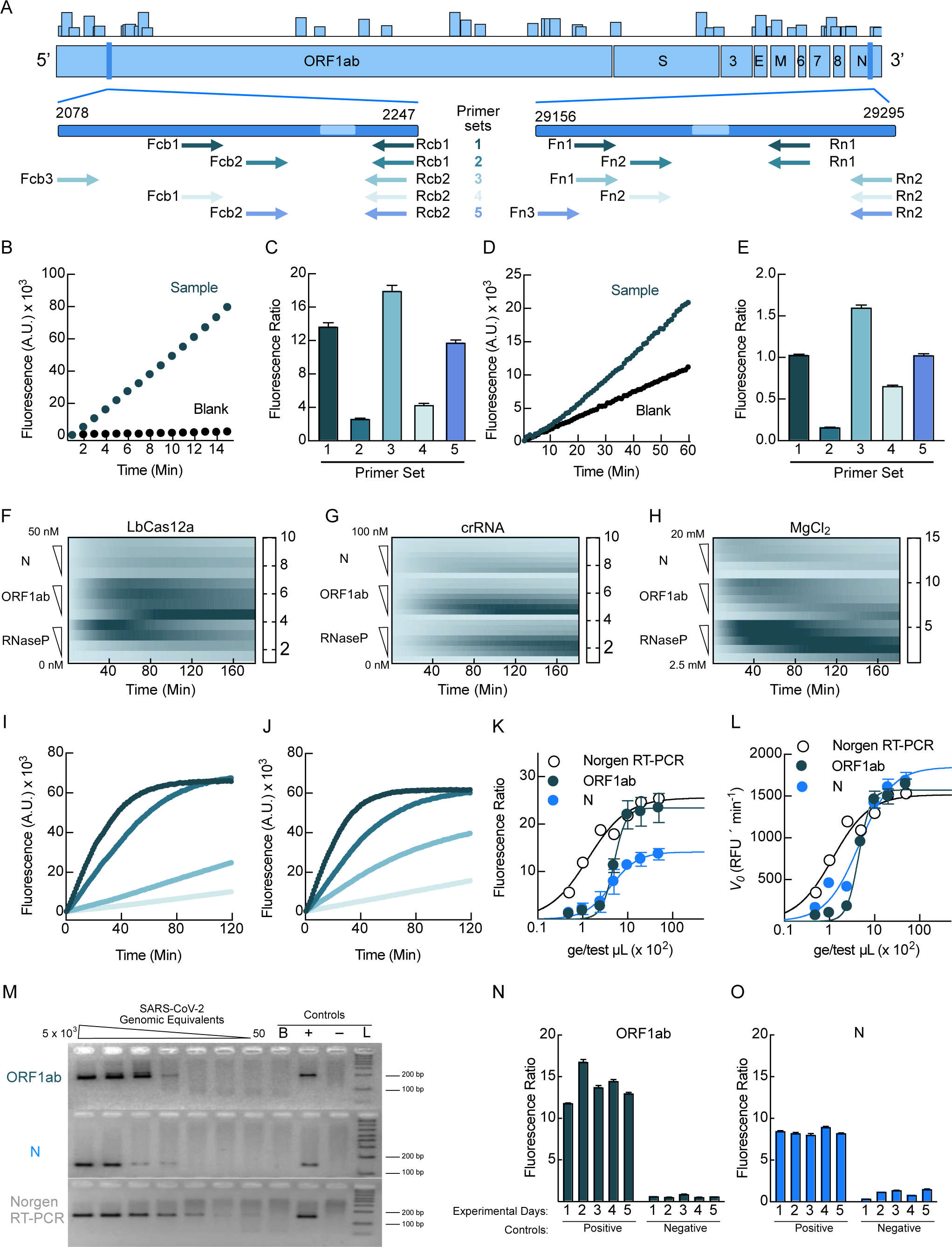
Optimization and analytical validation. (A) Schematic representation of the SARS-CoV-2 genome, primer localization for ORF1ab and N target genes, and reported frequencies of mutations (histogram above). (B) Example fluorescence time course of CRISPR/Cas12a-mediated recognition of ORF1ab. (C) Fluorescence ratios comparing five primer combinations for ORF1ab. Colors for the primer combinations are as in (A). (D) Same as panel (B) for the N detection *locus*. (E) Comparison of five primer combinations for the N detection region. (F-H) Heatmaps displaying the CRISPR/Cas12a reaction components that were optimized, namely LbCas12a (F), crRNA (G), and magnesium (H), for both viral detection *loci* and for the sample control human RNaseP. Reaction fluorescence ratios are depicted with continuous color shading. The concentrations of Cas12a, crRNA, and Magnesium were 10 nM, 15 nM, and 10 mM, respectively, unless it was the variable under study. (I) Time courses of the CRISPR/Cas12a-mediated detection of ORF1ab at increasing genome equivalents of SARS-CoV-2 RNA. (J) Same as panel (I) for the N *locus*. Genome equivalents ranged from 5 x 10^3^ to 5 x 10^1^ per reaction microliter. (K) Comparison of the fluorescence ratio as function of input genome equivalents of SARS-CoV-2 RNA in the RT-PCR reaction for ORF1ab and N *loci* and the commercial 2X One-Step RT-PCR Master mix from Norgen. (L) Initial velocity (V_0_) dependency of genome equivalents. (M) Gel electrophoresis analysis of RT-PCR products with varying genome equivalents for ORF1ab, N, and using the Norgen Biotech commercial one-step kit. Panels (N) and (O) show day-to-day reproducibility assays for ORF1ab and N *loci*, respectively. Fluorescence ratio is defined as the fluorescence of the test sample over that of the RT-PCR non-template control (blank) at a given time. Error bars represent the standard deviation of at least three independent measurements.

### RT-PCR optimization and primer selection

Most of molecular tools to detect SARS-CoV-2 use a combination of a reverse transcriptase and a DNA polymerase followed by result visualization using real-time thermocyclers, limiting LMICs. These countries could overcome such limitations with alternative readouts, i.e. from in-gel visualization or fluorescence-mediated direct observation under a transilluminator. Here, we describe the optimization of the reported conditions for M-MLV RT and Taq DNA polymerase as adapted from the open-source protocol BEARmix (Graham et al., 2021). RT-PCR reactions can be done in a single-step or two-step workflow. First, we compared the amplification performance using both enzymes in a single reaction *versus* adding Taq DNA polymerase once the RT step was completed, as differences in efficiency have been reported (Paula et al., 2004; Wacker and Godard, 2005). We used CRISPR/Cas12a as a method for comparing the amplification results using the fluorescence ratio between the test sample over the non-template control. No considerable differences were observed for samples with high viral load, whereas a fluorescence ratio greater than two was observed in low viral load samples for the one-step approach (Figure S3). Then, we evaluated the duration of the reverse transcription reaction. A 2.3-fold increase in the signal ratio was observed for samples with low viral load at 20 minutes of reverse transcription as compared to 10 minutes (Figure S3). Thus, a single step RT-PCR using 20 min of retro-transcription was suitable.

Based on the previously selected conditions we performed titrations for both enzymes. We found that final concentrations of 1.6 ng/µL of Taq DNA polymerase and 1.7 ng/µL of M-MLV RT were sufficient to achieve a fluorescence ratio ≥ 2 for both high and low viral RNA load samples (Figure S4). The viral RNA sample volume was evaluated to determine if the fluorescence signal in low viral load could be increased. However, no substantial differences were observed, and two microliters of sample volume in a 20 µL final reaction volume were used for the following standardization protocol (Figure S5). Finally, five primer sets for each detection *loci* were evaluated under the optimized RT-PCR amplification using a CRISPR/Cas12a assay (Figure 2A). The primer set 3 was selected for both detection *loci* as they showed the highest fluorescence ratio for the ORF1ab (Figures 2B, 2D) and the N targets (Figures 2C, 2E). Amplification with the selected primers generated 168-bp and 131-bp PCR products for ORF1ab and N, respectively. Strong fluorescence in the CRISPR/Cas detection reaction was observed for the ORF1ab target with a mean of 18-fold fluorescence ratio at 30 minutes (Figures 2B-C). The fluorescence ratio for the N target was 2-fold lower than that for ORF1ab. Likely due to an increased unspecific fluorescence signal for the non-template control (Figure 2D-E).

### Optimization of the CRISPR/Cas12a detection system

Several factors can influence the fluorescence readout in CRISPR/Cas detection assays, such as Cas:crRNA molar ratio, crRNA sequence, fluorophore quencher (FQ)-labeled single-stranded DNA (ssDNA) reporter probe, magnesium concentration, or temperature (Bai et al., 2019; Huang et al., 2020; Tsou et al., 2019). We evaluated three variables for the system based on reported conditions (Broughton et al., 2020), for both SARS-CoV-2 targets and the human RNaseP control. First, a range between 5 – 50 nM LbCas12a was titrated against 15 nM crRNA. This showed that 10 nM of Cas12a was sufficient for a fluorescence ratio > 3 for N and ORF1ab targets. Similarly, a fluorescence ratio of 2.5 was the case for the control target RNaseP with 10 nM of Cas12a (Figure 2F). As for the N target, higher Cas12a concentrations showed higher unspecific noise with the consequent reduction of the fluorescence ratio. On the other hand, for ORF1ab a higher concentration of Cas12a did not show a similar effect, the highest fluorescence ratio was found with 25 nM Cas12a at 30 minutes (Figure 2F). However, after 90 minutes of the reaction an increasing unspecific signal was observed for Cas12a concentrations starting at 5 nM. For the RNaseP target, higher Cas12a concentration increased the fluorescence ratio (Figure 2F).

Next, we tested crRNA concentration at a range between 5 – 100 nM against 10 nM LbCas12a (Figure 2G). For all targets, 15 nM crRNA resulted in the highest fluorescence ratio, more than twofold at 30 minutes. For all targets, crRNA concentrations of more than 25 nM showed increased unspecific backgrounds (Figure 2G). Also, a range between 2.5 – 20 mM MgCl_2_ was evaluated. For the N target, no considerable differences in the fluorescence ratio were observed between 10 – 20 mM MgCl_2_. On the other hand, for ORF1ab and RNaseP targets, the highest fluorescence ratios (> 10) were observed with 15 mM MgCl_2_ or higher at 30 minutes (Figure 2H). xFinally, the selected optimal conditions for sample testing were 10 nM LbCas12a, 15 nM crRNA and 15 mM MgCl_2_. Additionally, a synthetic dsDNA template for each target was used to estimate the analytical sensitivity of the selected conditions. A ten-fold serial dilution of each dsDNA template (10 nM to 1 pM) was evaluated, and a 1.5 fluorescence ratio was arbitrarily considered as a threshold value of detection. For the N target, 1 nM was the minimal concentration of molecules detected. In contrast, ORF1ab and RNaseP targets could be detected at 10 times lower concentration than N, allowing the detection of 100 pM (Figure S6).

### Analytical sensitivity estimation

To assess the detection limit of the optimized CRISPR-based detection system coupled to RT-PCR, we used purified SARS-CoV-2 genomic RNA (isolate USA-WA1/2020, obtained from BEI Resources (Cat # NR-52347)). For the ORF1ab target, high fluorescence ratios were observed for at least 5 x 10^2^ ge/µL at 30 minutes. Lower viral RNA copy numbers resulted in a drastic decrease of the fluorescence signal as observed by the reduced ratios towards 3-fold (Figure 2K). Likewise, detection of the N target was achieved up to 10^2^ ge/µL, differentiating from the blank reaction with a fluorescence ratio > 4 (Figure 2J). Our locally produced enzymes for RT-PCR and optimized conditions were compared to a commercial one-step RT-PCR kit (Norgen BioTek) using the N primer set (Figure 2K). Fluorescence ratios from the commercial kit were similar to the ratios for the ORF1ab target and 2-fold higher than the N target obtained with the produced enzymes (Figure 2K). This likely results from the 2-fold increase of V_0_ in the non-template control (RT-PCR blank) for the N target as compared to the ORF1ab. Thus, no apparent differences were observed when the initial velocity (V_0_) of the fluorescence time courses was used (Figure 2L).

An alternative to the fluorescence readout for LMICs is to use widely available gel electrophoresis. Gel visualization of the amplification products showed expected bands for ORF1ab (168 bp) and N (131 bp) targets up to 5 x 10^2^ ge/µL in the RT-PCR reaction. Unlike the commercial kit, visible bands were detected at 2.5 x 10^2^ ge/µL with weak bands up to 5 x 10^1^ ge/µL (Figure 2M). Finally, the reproducibility of the CRISPR/Cas12a method was evaluated along five different days using sample pools for positive and negative controls. Very limited day-by-day variation was observed (Figure 2N-O). Average fluorescence ratios of 14± 2 RFU and 0.6 ± 0.1 RFU were estimated for the positive and negative controls of the ORF1ab target, respectively. Likewise, fluorescence ratios of 8.4 ± 0.3 RFU and 1.1 ± 0.4 RFU were calculated for the positive and negative controls of the N target, respectively.

### Clinical Validation

Clinical evaluation of molecular methods used for SARS-CoV-2 detection have shown sensitivities ranging between 50% – 90% and a specificity near 99% (Stites and Wilen, 2020; Vandenberg et al., 2021; Wang et al., 2020). The diagnostic performance of our molecular toolkit was evaluated by testing clinical samples. One hundred clinical samples collected during 2020 were analysed by the gold-standard RT-qPCR test. Positive (N = 50) and negative (N =50) for SARS-CoV-2 were randomized and analyzed with the molecular toolkit developed here. Clinical samples showed varying fluorescence ratios, between 0.71 and 11.98 for the N gene, and 0.45 to 19.95 for the ORF1ab (Figure 3A, Figure S7). Using V_0_ as the outcome variable, positive samples presented a V_0_ between 226 ± 2 – 1695 ± 12 RFU × min^-1^ and 40 ± 1 – 1614 ± 13 RFU × min^-1^, for the N and ORF1ab *loci* respectively (Figure S7). Negative clinical samples resulted in low fluorescence ratios and V_0_ (Figure 3A, Figure S7). For all positive samples, fluorescence ratios and V_0_ were considerably higher than the negative controls while for the RNaseP sample control was broadly distributed (Figure S8). The fluorescence ratios and V_0_ for both N and ORF1ab targets showed a positive dependence on the viral RNA load of the samples (Figures 3B, C), while no correlation was observed with the respective RNaseP fluorescence ratios or V_0_ (Figures 3D,E).

**Figure 3:**
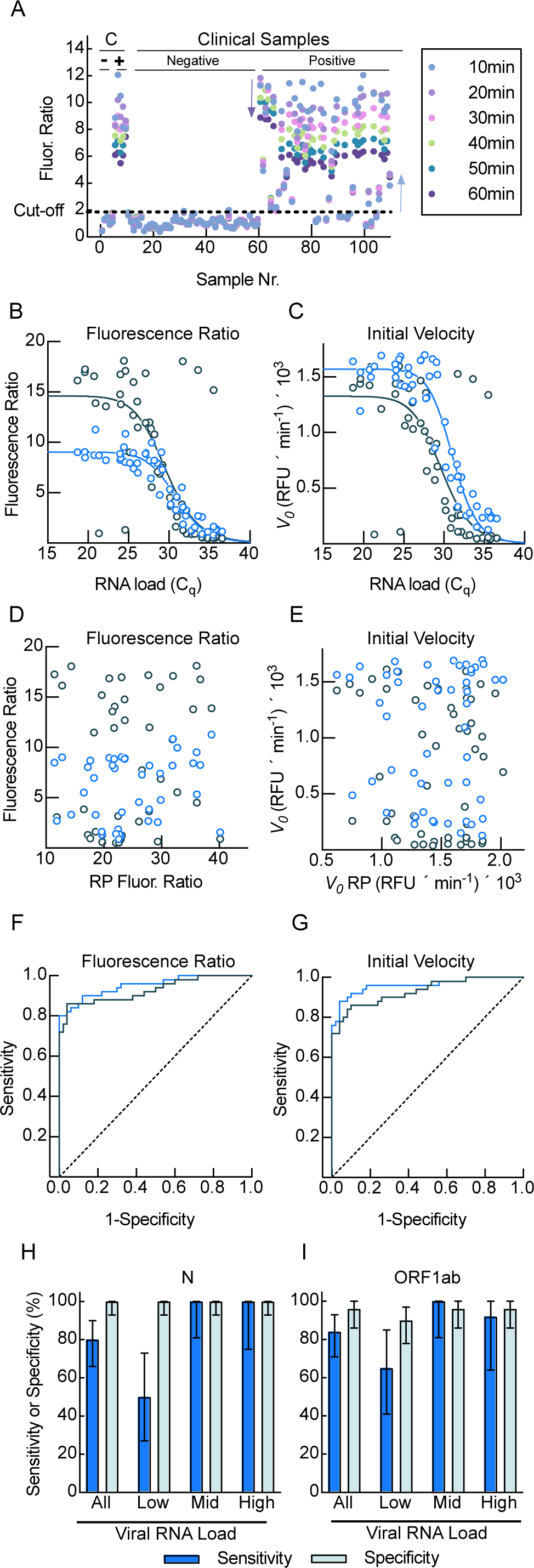
Test performance with clinical samples. (A) Distribution of fluorescence ratios for positive and negative controls (n = 10) and unknown samples (n = 100) for the N target. Dashed line indicates the threshold as calculated by the ROC analysis. (B) Fluorescence ratio as a function of viral RNA load (Cq values) for both SARS-CoV-2 targets, N (blue) and ORF1ab (green). (C) Same as panel (B) but using the initial velocity (V_0_). Fluorescence ratio (D) and initial velocity (E) dependence as a function of the RNaseP target. Colours as in (B). ROC curve based on fluorescence ratio (F) or initial velocity (G). ROC curves were obtained independently for each evaluated SARS-CoV-2 target; colours are as in (B). (H) N target detection sensitivity and specificity with 95% CI for all samples and by viral RNA load. (I) As H for ORF1ab target.

Thus, the performance of the molecular toolkit is dependent on the viral RNA load rather than on variations during sample collection or RNA extraction.

The detection boundaries were estimated by calculating a ROC curve from the fluorescence ratio and V_0_ for both, N and ORF1ab regions (Figures 3F, G). No significant difference was observed between the ROC curves by either analytical approach (P = 0.508). The sensitivity and specificity using the fluorescence ratio and V_0_ data analysis were determined for all samples and within groups based on viral RNA load (Figures 3H,I, Tables S1-S3). For the N target, the overall sensitivity and specificity, as calculated using fluorescence ratios and a cut-off of 1.86, were 80% and 100%, respectively. If V_0_ was used, the analysis showed 88% sensitivity and 96% specificity (cut-off value of 253 RFU × min^-1^) (Figure 3H, Tables S1). The samples that were misclassified by the fluorescence ratio threshold showed a Cq value > 33 and V_0_ < 300 RFU × min^-1^. However, two of the correctly classified samples presented Cq values of 34 but V_0_ > 330 RFU × min^-1^. On the other hand, for the ORF1ab target, the fluorescence ratio analysis (cut-off value of 0.86) showed 84% sensitivity and 96% specificity, whereas the V_0_ analysis (cut-off value of 73 RFU × min^-1^) showed a sensitivity of 86% and a specificity of 90% (Figure 3I, Tables S1). Similarly to N target detection, false negative results presented a Cq value > 33 but with V_0_ < 450 RFU × min^-1^. Nonetheless, five true positive results showed a Cq > 33 with V_0_ ranging between 226 and 277 RFU × min^-1^. With regard to the false positive results reported herein, a V_0_ < 360 RFU × min^-1^ was estimated (Cq values not determined as they were negative with the gold standard). Visual detection by gel electrophoresis showed lower sensitivity values for both N (66%) and ORF1ab (50%) targets, but similar specificity to fluorescence readout (Tables S1). All samples were divided by RNA load using the Cq values, high (<25), medium (>28<), and low (>31). Our results show an increased sensitivity for both targets in samples with medium to high viral RNA loads, achieving almost 100% sensitivity. The increase was independent of the analysis used, either the fluorescence ratios or V_0_ analysis (Tables S2). However, samples with low viral RNA load showed lower sensitivity, less than 70% (Figure 3H,I, Tables S2).

## Discussion

A prompt epidemiological surveillance of infectious diseases requires decentralized detection capacity, acquired by enabling direct detection of the pathogenic entity in primary health care and available laboratories. Although many alternative methods have been described, RT-PCR remains the gold standard for detection of SARS-CoV-2 and other respiratory viruses (Beck and Henrickson, 2010; Benzigar et al., 2020; Böger et al., 2021; Lin et al., 2020). Furthermore, PCR and qPCR methods are a straightforward analytical solution for a number of emerging pathogens. For instance, they were rapidly deployed in recent virus outbreaks including Influenza, SARS and MERS, Zika, Dengue, and Chikungunya, among others, that pose a serious threat to humans, either because of their pandemic potential or due to their recurrent appearance affecting severely communities in LMICs (Johani and Hajeer, 2016; Johnson et al., 2016; Ravina et al., 2021; Silva et al., 2019). Scientists and health professionals promptly developed solutions to cope with the lack of molecular detection platforms. Thus, LMICs have the theoretical and practical bases to rapidly design, validate, and use PCR-based methods for any pathogenic threat. Yet, the Covid-19 pandemic unmasked an unexpected complication–a shortage of laboratory testing supplies due to an unprecedented global demand. Paradoxically, globalization hampered LMICs for accessing molecular detection solutions, yet unlocked the access to information for driving local efforts to deal with the global market limitations. The latter is further evidenced by several and remarkable open access initiatives like BEARmix (Graham et al., 2021), Addgene (https://www.addgene.org/), BEI Resources (https://www.beiresources.org/), Nextstrain (https://nextstrain.org/), and GISAID (https://www.gisaid.org/). The work presented here was fully performed in a LMIC laboratory and used all available open access resources to implement a molecular toolkit aiming to provide versatile solutions for LMICs, and to cope with the international shortage of testing supplies for SARS-CoV-2 or any other pathogenic agent.

The molecular toolkit validated here shows high versatility to be adapted to different contexts by exploiting widely accepted and known PCR-based methods (Figure 4A). The initial stage focused on providing a minimal set of essential recombinant enzymes that allows the amplification and visualization of viral genes in laboratories with minimal equipment. The open access BEARmix initiative was particularly important as they kindly provided plasmids, protocols, and initial conditions along their development and prior to any publication (Graham et al., 2021). Yet, some routines of the protein production workflow needed to be adapted due to equipment availability. For instance, enzyme production can be achieved to high purity in laboratories lacking a FPLC instrument by using salt gradients set up manually and inexpensive plastic syringes. This, in turn, allows any laboratory with minimal *E. coli* cultivation capacities to produce high purity enzymes. Other simpler methods have been also reported and could be tested. Bhadra *et al*. used crude heat-treated extracts as a source of Taq, Phusion and Bst DNA polymerases, and M-MLV RT enzymes with promising results (Bhadra et al., 2018). In addition to RT-PCR enzymes we report the production of LbCas12a, also in a minimal laboratory setup. Altogether, the proposed four day/person workflow allows the production of enzymes for over fifty thousand tests. Further scaling or implementation in multiple laboratories could satisfy the testing needs of an entire city or country.

**Figure 4.**
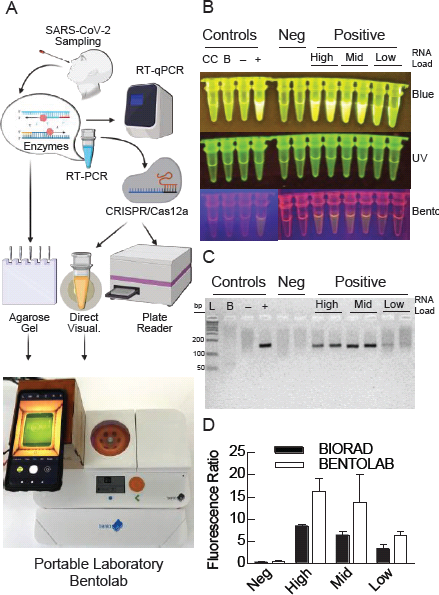
Alternative readouts and portability of molecular detection with BentoLab. (A) Scheme of the different analytical options for the method reported here. (B-D) Two clinical samples of each RNA load group (low, medium and high), and two negative samples are shown were analyzed using the N target. (B) Fluorescent signals visualized in tubes using three different transilluminators: blue light (470 nm) (top), UV (middle), and BentoLab (bottom). Controls included the CRISPR control for background fluorescence (CC), the RT-PCR non template control (blank). (B), and pools of negative or positive samples. (C) Agarose gel (5%) electrophoresis of conventional RT-PCR amplified products. L, DNA ladder; B, a PCR blank reaction (no template); plus and minus signs indicate positive and negative controls, respectively. (D) Ratio of CRISPR-Cas12a fluorescent signals of the test sample to that RT-PCR non-template control. Error bars indicate standard errors obtained from duplicate experiments.

The focus on RT-PCR methods unlocks several alternatives to visualize the presence or absence of a target genetic material in a patient sample (Figure 4A). Such versatility is further enhanced by coupling RT-PCR to detection mediated by CRISPR-Cas12a. In this context, the production of crRNA molecules from DNA templates can be easily performed to satisfy the demand (Figure S1). Additionally, CRISPR-Cas12a can be used in both, qualitative or quantitative detection, ways. The direct visualization of fluorescence in tubes allows any laboratory using a blue light transilluminator to qualitatively assign the presence or absence of amplified genetic material (Figure 4B) (Chen et al., 2020; Ding et al., 2020; Huang et al., 2020; Pang et al., 2020; Xiong et al., 2020). Our system is capable of detecting an observable signal after only 15 minutes of the CRISPR-Cas12a reaction (Figure S9). Another low-cost method uses widely available gel electrophoresis systems. Albeit the sensitivity is lower than fluorescence-based methods (Figure S10, Tables S2), agarose gel electrophoresis offers an alternative for laboratories equipped with only a thermocycler and transilluminator. Importantly, the specificity by agarose gel electrophoresis is not compromised and the sensitivity for high viral RNA load samples is still competitive (Figure S10). Albeit not tested here, Graham *et al*. used TaqMan probes for direct visualization of the genetic material using transilluminators with high sensitivity (Graham et al., 2021). Thus, also in the absence of LbCas12a, the molecular toolkit could be coupled to direct visualization of test results in laboratories with minimal molecular biology facilities.

Yet, the demand for molecular testing can exponentially escalate as the pandemic expands to poorly accessible areas. For instance, Peru holds the highest death toll per million of habitants, a country characterized by social and economic disparities if comparing cities to small towns. The latter still lack accessibility to molecular testing, even after more than a year into the Covid-19 pandemic. There are several reasons that can account for such shortages in LMICs, a prominent one being the lack of any laboratory infrastructure in remote rural cities, towns, and communities. To cope with these limitations, we applied the molecular toolkit established here using the portable laboratory from BentoLab that consists of a thermocycler for 32 samples, a microcentrifuge, a gel electrophoresis chamber, and a small transilluminator (Figure 4A). We show that the BentoLab is capable of amplifying SARS-CoV-2 genetic material (Figure 4B) and allows the visualization of results with our molecular toolkit (Figures 4B, 4C). Furthermore, a comparison of the amplification of target RNA between the BentoLab and a regular thermocycler by quantitatively measuring Cas12a-dependent fluorescence shows that the portable laboratory can be a solid alternative (Figure 4D). Thus, the molecular toolkit described here can potentially unlock the molecular detection of pathogenic agents even in resource-limited regions.

Alternatively, fluorescence plate readers are common in different laboratory setups and could be used for quantitative detection of SARS-CoV-2 (Figures 2, 3)(Ding et al., 2020; Ganguli et al., 2020; Sun et al., 2021; Woo et al., 2020; Xiong et al., 2020). Our toolkit used clinical samples to determine critical detection parameters, showing competitive sensitivity and specificity (Figure 3). Although the specificity was high regardless of viral RNA load, the sensitivity decreased with lower viral RNA load (Figure 3B,C). Thus, low viral loads may result in false negative results. Yet, molecular detection is one variable that is considered to assess the likelihood of infection that a person may hold. Indeed, most diagnostic algorithms also consider symptoms, comorbidities, vicinity to diagnosed contacts, prevalence of SARS-CoV-2 in the population being tested, among other factors. Thus, even low sensitivity in detection systems can greatly contribute for better diagnostic assessments at the clinical level. Our system shows an overall sensitivity of 86% for samples of unknown viral load and increases to 100% for patients with medium to high viral load. A recent study found that Cq values greatly varied among SARS-CoV-2-infected people, with a distribution peaking at Cq values of 23-25 (Buchan et al., 2020). In that context, our system should be able to correctly detect all analyzed samples.

Altogether, the presented study reports a molecular toolkit that can be produced and used in a variety of laboratory setups. Furthermore, the BentoLab portable laboratory could unlock molecular detection even in low-resource settings lacking any laboratory infrastructure. Poor infrastructure combined with reduced accessibility to reagents could be overcome in LMICs. To our knowledge, this is the first report of a molecular detection platform entirely produced in a LMIC laboratory setup, providing detailed methods and final protocols to be used, and that can be adapted for any forthcoming pathogenic threat.

## Data Availability

All data is available in the manuscript or its supplemental information

## Acknowledgments

We are very thankful to Dr. Marcos Milla for donating equipment that was used in this study. We would also like to thank all lab members of the Del Valle, Adaui, and Milón groups for their help, support, helpful discussions, and great working atmosphere. This work was supported by grants from the Peruvian *Fondo Nacional de Desarrollo Científico, Tecnológico y de Innovación Tecnológica* [070-2020-FONDECYT] to VA and [036 -2019-FONDECYT-BM-INC.INV] to PM. Funding for open access was provided by the Universidad Peruana de Ciencias Aplicadas.

## Author Contributions

VA, KP, RA, and PM conceived the project. RA, KP, JAN, GM, and JML performed experiments. RA, JDV, KP, JAN, and PM analysed the data. RA, KP, VA, and PM wrote the manuscript with the input of all authors.

## Star Methods

### Resource availability

**Lead contact:** Materials and information can be asked to Pohl Milón

### Key Resources Table

Uploaded separately

### Materials availability

The sequences of all generated materials are provided in the article or in Supplemental Information.

### Data and code availability

The published article includes all datasets generated or analyzed during this study. This study did not generate/analyze code.

### Experimental model and subject details

#### Microbes

*E.coli* BL21(DE3)pLysS cells were cultivated in Luria - Bertani medium.

### Methods

#### Protein expression and purification

Plasmids coding for M-MLV reverse transcriptase (RT) and Taq DNA polymerase, as part of the BEARmix project, were obtained from the Tjian-Darzacq laboratory (https://gitlab.com/tjian-darzacq-lab/bearmix) (Graham et al., 2021). Both proteins are fused with an N-terminal His-tag. Plasmids coding for full-length LbCas12a (#113431) (Chen et al., 2018) and TEV protease S219V (#8827) (Kapust et al., 2001) were obtained from Addgene. Both LbCas12a and TEV protease are fused with an N-terminal His-tag, maltose binding protein (MBP) and TEV protease cleavage site. Plasmids were transformed independently into chemically competent BL21(DE3) *E. coli* using the Mix&Go *E. coli* transformation kit (Zymo Research).

For the M-MLV RT, a single colony was inoculated into Luria-Bertani (LB) medium supplemented with 30 µg/mL kanamycin (Kan) and 30 µg/mL chloramphenicol (Cam) and incubated at 37°C and 150 rpm overnight. The starter culture was subsequently used to inoculate fresh LB medium (+ 30 µg/ml Kan and 30 µg/ml Cam) cultures to an optical density of 0.05 measured at a wavelength of 600 nm (OD600 = 0.05) and grown at 37 °C and 150 rpm until cells reached OD600 = 0.8. Protein expression was induced with 1 mM isopropyl β-d-1-thiogalactopyranoside (IPTG) at 37 °C for 3 h with agitation at 150 rpm. Cell pellets were harvested and washed with TAKM buffer (50 mM Tris-HCl pH 7.4, 70 mM NH4Ac2, 30 mM KCl, 10 mM MgCl_2_) by centrifugation at 5 000 rpm and 4 °C for 5 min. After that, cell pellets were harvested again and resuspended in MB1 (50 mM Tris-HCl pH 8, 100 mM NaCl, 10 mM imidazole, 6 mM β-mercaptoethanol, 0.1% Tween 20) with 100 µg/mL lysozyme, 40 µg/mL DNase I, 10 mM MgCl_2_ + Complete protease inhibitor (Roche) and stored at -80 °C.

Cells were lysed by two freeze/thaw cycles and NaCl concentration was fixed to 370 mM. The lysate was centrifuged at 10 000 rpm and 4 °C for 45 min. The supernatant was filtered through a 0.45-μm Millex-HV syringe filter (Sigma-Aldrich) and manually loaded onto a 1 mL HisTrap FF Crude column, pre-equilibrated with MB1. The column was washed twice with MB2 (50 mM Tris-HCl pH 8, 500 mM NaCl, 10 mM imidazole, 6 mM β-mercaptoethanol, 0.1% Tween 20) and MB1. The protein was eluted with 250 mM imidazole in MB3 (50 mM Tris-HCl pH 8, 100 mM NaCl, 250 mM imidazole, 6 mM β-mercaptoethanol, 0.1% Tween 20). Fractions were analyzed by 10% SDS-PAGE and the relevant fractions containing protein were pooled and dialyzed overnight at 4 °C against MB4 (50 mM Tris HCl pH 8, 100 mM NaCl, 0.1 mM EDTA, 6 mM β-mercaptoethanol, 0.1% Tween 20). The protein was loaded onto a 1 mL HiTrap SP column pre-equilibrated with MB4 and washed twice with the same buffer. M-MLV was eluted with linear gradient using MB4 (100 mM NaCl) and MB5 (50 mM Tris-HCl pH 8, 1 M NaCl, 0.1 mM EDTA, 6 mM β-mercaptoethanol, 0.1% Tween 20) in 20 CVs, analyzed by 10% SDS-PAGE, pooled and dialyzed in MB6 (50 mM Tris HCl pH 8, 100 mM NaCl, 0.1 mM EDTA, 1 mM DTT (added just before use), 0.1% Tween 20 or Triton X-100, 50% glycerol).

For the Taq DNA polymerase, cellular expression was done as described before for M-MLV RT. Cell pellets were also harvested and washed with TAKM buffer. After that, cell pellets were stored resuspended in TaB1 (50 mM Tris HCl pH 8, 500 mM NaCl, 0.1% NP-40 and 0.1% Triton X-100) with 100 µg/mL lysozyme, 40 µg/mL DNase I, 10 mM MgCl_2_ + Complete protease inhibitor (Roche) at -80 °C. Cells were lysed by two freeze/thaw cycles and heated at 80 °C for one hour. The lysate was centrifuged at 10 000 rpm and 4 °C for 45 min and the buffer was fixed to 6 mM β-mercaptoethanol, 10 mM imidazole, 10% glycerol. The supernatant was filtered and manually loaded onto a 1 mL HisTrap FF Crude column that was pre-equilibrated with TaB1. The column was washed with TaB2 (50 mM Tris HCl pH 8, 500 mM NaCl, 0.05% NP-40, 5% glycerol, 10 mM imidazole, 6 mM β-mercaptoethanol) and TaB3 (50 mM Tris HCl pH 8, 100 mM NaCl, 0.05% NP-40, 5% glycerol, 10 mM imidazole, 6 mM β-mercaptoethanol), and eluted with TaB4 (TaB3 buffer containing 300 mM imidazole). Fractions were analyzed by 10% SDS-PAGE and relevant fractions containing the protein were pooled and dialyzed overnight at 4 °C against TaB5 (50 mM Tris HCl pH 8, 100 mM NaCl, 0.05% NP-40, 10% glycerol, 6 mM β-mercaptoethanol). Pooled fractions were loaded onto a 1 mL HiTrap Heparin HP column and washed twice with TaB5. Fractions were eluted with a salt gradient up to 1 M NaCl with TaB6 (TaB5 containing 1 M NaCl), analyzed by 10% SDS-PAGE, pooled and dialyzed in TaB7 (50 mM Tris HCl pH 8, 100 mM NaCl, 0.1 mM EDTA, 1 mM DTT, 50% glycerol).

For the TEV protease, a single colony was inoculated into LB medium supplemented with 100 µg/mL ampicillin (Amp) and 30 µg/mL (Cam) and incubated at 37 °C and 150 rpm overnight. The starter culture was subsequently used to inoculate fresh LB medium (+ 100 µg/mL Amp and 30 µg/mL Cam) cultures to an OD600 = 0.05 and grown at 37°C and 150 rpm until cells reached OD600 = 0.5. Protein expression was induced with 1 mM IPTG at 30 °C and 150 rpm for 3 h. Cell pellets were harvested by centrifugation at 5 000 rpm and 4 °C for 10 min and resuspended in HAKM10 buffer (50 mM HEPES pH 7.4, 70 mM NH4AC, 50 mM KCl, 10 mM MgCl_2_, 6 mM β-mercaptoethanol). Then, 100 µg/mL lysozyme and Complete Protease Inhibitor Cocktail (1 tablet/25 mL) (Roche) were added to the cell suspension, mixed and stored at -80 °C. Cells were lysed by two freeze/thaw cycles in the presence of 40 µg/mL DNase I. The lysate was centrifuged at 10 000 rpm and 4 °C for 45 min. The supernatant was filtered through a 0.45-μm Millex-HV syringe filter (Sigma-Aldrich) and manually loaded onto a 1 mL HisTrap FF Crude column that was pre-equilibrated with TeB2 (20 mM HEPES pH 7.4, 300 mM NaCl, 6 mM β-mercaptoethanol containing 10 mM imidazole). The column was washed with TeB3 (50 mM HEPES pH 7.4, 300 mM NaCl, 50 mM imidazol, 6 mM β-mercaptoethanol) and eluted with TeB4 (50 mM HEPES pH 7.4, 300 mM NaCl, 100 mM imidazol, 6 mM, 6 mM β-mercaptoethanol). Eluted fractions were analyzed by 12.5% SDS-PAGE and Coomassie blue staining. Fractions containing the TEV protease were pooled and dialyzed overnight at 4 °C against TeB5 (25 mM HEPES pH 8.0, 200 mM NaCl, 20% glycerol, 1 mM ethylenediaminetetraacetic acid (EDTA), 6 mM β-mercaptoethanol) using a D-Tube™ Dialyzer Maxi, MWCO 12-14 kDa (Sigma-Aldrich).

For LbCas12a, a single colony was inoculated into LB medium supplemented with 100 µg/mL Amp and incubated at 37°C and 150 rpm overnight. The culture was diluted in fresh LB medium (+ 100 µg/mL Amp) and grown at 37 °C and 150 rpm until OD600 of 0.5 was reached. The cultures were incubated on ice for 15 min before induction with 0.2 mM IPTG at room temperature and 150 rpm overnight. Cell pellets were harvested by centrifugation at 5000 rpm and 4 °C for 10 min, resuspended in HAKM buffer with 400 mM NaCl, 100 μg/ml lysozyme, 40 µg/mL DNase I + Complete protease inhibitor (Roche) and stored at -80 °C. The cell pellet was lysed by 2 freeze/thaw cycles. The lysate was centrifuged at 10000 rpm and 4 °C for 45 min. The supernatant was filtered through a 0.45-μm Millex-HV syringe filter (Sigma-Aldrich) and loaded onto a 1 mL HisTrap FF Crude column that was pre-equilibrated with CB1 buffer (20 mM HEPES pH 7.4, 300 mM NaCl, 10 mM Imidazole, 6 mM β-mercaptoethanol). The protein was washed with CB2 (50 mM HEPES pH 8, 300 mM NaCl, 50 mM imidazole, 6 mM β-mercaptoethanol) and CB3 (50 mM HEPES pH 8, 300 mM NaCl, 100 mM imidazole, 6 mM β-mercaptoethanol) and eluted with CB4 (50 mM HEPES pH 8, 300 mM NaCl, 250 mM imidazole, 6 mM β-mercaptoethanol). Eluted fractions were analyzed by 7.5% SDS-PAGE and Coomassie blue staining. Fractions containing LbCas12a were pooled and TEV protease was added to 2.21 μM final concentration. The sample was dialyzed overnight at 4 °C against CB5 (25 mM HEPES pH 7.5, 200 mM NaCl, 20% glycerol, 1 mM EDTA, 6 mM β-mercaptoethanol) using a D-Tube™ Dialyzer Maxi, MWCO 12-14 kDa (Sigma-Aldrich).

For further purification, the dialyzed sample was manually loaded onto a 1 mL MBPTrap HP column that was pre-equilibrated with CB6 (20 mM HEPES pH 7.5, 125 mM KCl, 5% glycerol, 1 mM β-mercaptoethanol). The flow through (FT) with the protein was collected and the remaining MBP trapped in the column was washed with CB7 (20 mM Hepes pH 7.5, 125 mM KCl, 5% glycerol, 6 mM β-mercaptoethanol, 10 mM maltose), CB6 and UP water. Following, the FT was loaded in a 1 mL HiTrap SP HP column pre-equilibrated with CB6. LbCas12a was washed with CB6 and eluted with CB8 buffer (20 mM Tris-HCl pH 7.5, 200 mM NaCl, 20% glycerol, 1 mM DTT, KCl linear gradient). Recollected fractions were visualized by 7.5% SDS-PAGE and Coomassie blue staining. Fractions containing the LbCas12a protein were pooled and concentrated with an Amicon Ultra-4, membrane PLQK Ultracel-PL, 50 kDa (Sigma-Aldrich). The sample was then dialyzed overnight at 4°C against CB8 using a D-Tube™ Dialyzer Maxi, MWCO 12-14 kDa (Sigma-Aldrich).

Dialyzed proteins, M-MLV RT, Taq DNA polymerase and TEV protease were aliquoted and stored at -20 °C, LbCas12a was store at -80°C. Protein concentrations were determined by the Bradford assay using bovine serum albumin (BSA) for the standard curve.

#### crRNAs preparation

crRNAs were prepared from synthetic dsDNA templates (100 pmoles) using TranscriptAid T7 High Yield Transcription Kit (Thermo Fisher Scientific) at 37 °C for 3 h. Immediately thereafter, crRNAs were purified using Direct-zol RNA miniprep (Zymo Research) with a DNase I (30 U) digestion in a column step for 15 min at room temperature and eluted in nuclease-free water. Finally, crRNAs were quantified by NanoDrop One microvolume UV-Vis spectrophotometer (Thermo Fisher Scientific), aliquoted to working volumes and stored at -80 °C.

#### RT-PCR assays

The RT-PCR assay was standardized using pooled patient samples with high viral load (Cq < 30) and low viral load (Cq > 31), and non-template reactions as negative controls. Samples pools were prepared by mixing 10 high viral load and 10 low viral load samples, respectively. Initial conditions were based on reported data (Graham et al., 2021). Master mix reactions were prepared using the enzymes M-MLV RT and Taq

DNA polymerase, produced as described above. RT-PCR products were visualized in 5% agarose gel as described above.

In order to find optimal conditions, one-step and two-step RT-PCR reactions were compared. Additionally, reverse transcription time (10 and 20 minutes), different sample volumes (0.5, 1, 2, and 4 µL), different concentrations of Taq DNA polymerase (0.8 ng/µL, 1.6 ng/µL, 3.2 ng/µL, 8 ng/µL, and 16 ng/µL) and of M-MLV RT (0.8 ng/µL, 1.7 ng/µL, 3.4 ng/µL, and 5.1 ng/µL) were evaluated. Enzyme dilutions were prepared in MB6 buffer. The analytical sensitivity was evaluated using a range of SARS-CoV-2 genomic RNA (NR-52347, BEI Resources) between 5 x 10^3^ and 5 x 10^1^ viral genome equivalents (ge) per microliter of reaction. Additionally, the analytical sensitivity of the RT-PCR was compared with that of a commercial 2X One-Step RT-PCR Master Mix (Norgen BioTek) for the N target. The amplified products were verified by 5% agarose gel electrophoresis (TBE 1X, 70V for 45 minutes), and visualized using Safe-Green (N° Cat. G108-G, abm) in the loading buffer TriTrack DNA loading dye 6X (N° Cat. R1161, ThermoFisher) in a safeVIEW: LED/Blue Light transilluminator (Cleaver Scientific).

The optimized conditions of SARS-CoV-2 RNA amplification by RT-PCR used a 20 µL RT-PCR reaction with 1.6 ng/µL of Taq DNA polymerase, 1.7 ng/µL M-MLV RT, 0.4 mM dNTPs and 0.2 µM of each primer in RPB1X (50 mM Tris-HCl, 75 mM KCl, 3 mM MgCl_2_, 10% trehalose, 10 mM DTT, 0.1 mM EDTA, pH 8.4, 25 °C) with 2 µL of RNA sample. The reaction used a reverse transcription step at 50 °C for 20 minutes, followed by an initial denaturation at 95 °C for 5 minutes, and 45 cycles of denaturation at 95 °C for 3 seconds and a single annealing-extension step at 55 °C for 30 seconds.

#### CRISPR-Cas12a *trans*-cleavage assays

The CRISPR-Cas12a-based assays were standardized using synthetic dsDNA as templates in 96-well flat clear bottom black polystyrene tissue culture-treated microplates (Corning). Reporter probe (/56FAM/TTATT/3IABkFQ/, Macrogen) (Chen et al., 2018), recombinantly purified LbCas12a and *in vitro* transcribed crRNAs, prepared as described above, were used in all assays. CrB1X buffer (50 mM NaCl, 10 mM Tris-HCl, 100 µg/ml BSA, pH 7.9, 25 °C) + 15 mM MgCl_2_ was used for all steps, unless differently specified in the text or figure legends.

Initial settings were based on previous reports (Broughton et al., 2020; Chen et al., 2018). For each test, the crRNA was first refolded at 65 °C for 10 min in a heating block followed by slow cooling for 10 minutes at room temperature to enhance homogeneous folding. Afterward, the Cas12a:crRNA:reporter complex was formed for 10 min at room temperature. Each CRISPR-Cas12a assay consisted of 10 nM Cas12a and 15 nM crRNA using 0.3 µM reporter. Different concentrations of Cas12a (5 nM, 10 nM, 25 nM and 50 nM), crRNA (5 nM, 10 nM, 15 nM, 25 nM, 50 nM, 75 nM, 100 nM), and MgCl_2_ (2.5 mM, 5 mM, 10 mM, 15 mM, 20 mM) were tested to find optimal conditions. The analytical sensitivity was evaluated using a range of synthetic dsDNA template concentrations between 1 pM and 10 nM. Standardization assays were performed independently for both SARS-CoV-2 N and ORF1ab targets, and the human RNase P control template.

The optimized conditions of CRISPR/Cas12a assays used five microliters of unpurified RT-PCR products diluted in 85 µL of CrB1X (50 mM NaCl, 10 mM Tris-HCl, 100 µg/ml BSA, pH 7.9, 25 °C) + 15 mM MgCl_2_. Briefly, the Cas12a:crRNA:reporter complex was prepared at 10X concentration (100 nM Cas12a, 150 nM crRNA, 3 µM reporter probe) in CrB1X. Ten microliters of the 10X Cas12a:crRNA:reporter complex were mixed with 90 µL of the diluted RT-PCR products in a final volume of 100 µL to start the reaction. Kinetic assays monitored fluorescence signal at an excitation wavelength of 491±9 nm and an emission wavelength of 525±24 nm every minute for 2 hours at room temperature using a fluorescence plate reader (Cytation™ 5, BioTek Instruments). Fluorescence ratios (test sample fluorescence over that of CC) are used for analysis. Initial velocity (V_0_) was estimated using linear regression of the fluorescence time courses.

#### End-point CRISPR-Cas12a assays

For end-point CRISPR-Cas12a assays, SARS-CoV-2 N and ORF1ab targets and the human RNAseP control were amplified from clinical samples with confirmed SARS-CoV-2 infection status. Two clinical samples each with high, medium and low viral RNA load, as well as two negative (SARS-CoV-2 not detected) samples were evaluated using a conventional T-100 thermal cycler (Biorad) and the mobile molecular equipment BentoLab (https://www.bento.bio/). Thermocycler and gel electrophoresis/transilluminator modules of the portable BentoLab were used for RT-PCR, agarose gel visualization and CRISPR-Cas12a-based direct tube detection. RT-PCR and CRISPR-Cas12a reactions were performed as previously described. The fluorescence readout was monitored by naked-eye visualization every five minutes during one hour in a safeVIEW: LED/Blue Light transilluminator (Cleaver Scientific) and the electrophoresis/transilluminator module of the portable lab using a Black Box adaptation.

#### Clinical samples and validation

One hundred nasopharyngeal swabs of hospitalized patients during the COVID-19 pandemic in 2020 were used. Patients were categorized with mild or moderate disease as defined by Peruvian national guidelines. The study protocol was approved by the Research Ethics Board of the Instituto de Investigación Nutricional, Lima, Peru (N° 395-2020/CIEI-IIN) and by the Ethics Committee of the Universidad Peruana de Ciencias Aplicadas (N° FCS-CEI/187-07-20). The samples were obtained in the context of the epidemiological surveillance program according to the health directives of the National Center for Epidemiology, Disease Control and Prevention of the Ministry of Health of Peru. All methods were performed in accordance with the relevant guidelines and regulations. Therefore, the need of an explicit informed consent was waived by the Research Ethics Board of the Instituto de Investigación Nutricional, Lima, Peru Nasopharyngeal swabs samples were collected using COPAN swabs and UTM medium. The viral genetic material was extracted used the QIAamp viral RNA Mini Kit (250) (Cat N° 52906, QIAGEN) to perform the RNA extraction from 200 µL of the samples, according to the manufacturer’s instructions. Viral RNA obtained after the extraction was eluted in 100 µl of nuclease-free water. For this study, a double-blind approach was used with 50 positive samples SARS-CoV-2 and 50 negative samples SARS-CoV-2. The gold standard detection used the amplification of SARS-CoV-using Real Time ready RNA Virus Master and 250 nM specific primers for SARS-CoV-2:

Gene E, Gene RdRP (RNA-dependent RNA polymerase) and Gene N (Roche Diagnostic). The PCR condition was performed according to the manufacturer’s instructions. All procedures were performed using the Cobas Z480 System (Roche Diagnostic).The reference RT-qPCR test provided the quantification cycle (Cq, equivalent to the cycle threshold (Ct)), which are a semi-quantitative measure for the viral load, meaning that a low Cq-values indicate high viral load, and a high Cq-value indicate a low viral load. Cq values for the RdRP gene were used to assign RNA load categories.

#### Bioinformatic analyses, data processing, and statistical analysis

Ninety-six SARS-CoV-2 genomes (as of March 22, 2020) were aligned locally using the ClustalX package for Ubuntu with default parameters (File S1) (Larkin et al., 2007). Conserved regions were identified with the AliView software (Larsson, 2014). Protospacer adjacent motif (PAM) sequences (TTTV) for LbCas12a were searched to identify recognition 20nt-long sites. Double-stranded DNA (dsDNA) templates for crRNA generation through *in vitro* transcription were designed with a T7 promoter, followed by the scaffold and the specific recognition site (File S1). Synthetic dsDNA templates containing the recognition site and flanking regions were designed manually. Primers were designed manually or with the Primer3Plus v.2.4.2 server with the default settings and an average Tm of 56 °C (Untergasser et al., 2012).

Data analysis was performed using Prism 9 (GraphPad Software) and STATA v14.0 (StataCorp). Fluorescence signal over time was analyzed by linear regression fitting to determine the initial velocity at varying times (5-30 min). Fluorescence intensity ratios between synthetic template (cognate) and synthetic control (non-cognate) were calculated for each fluorescence reading during CRISPR-Cas12a optimization assays. Similarly, fluorescence intensity ratios between the test sample and RT-PCR non-template control (blank) were calculated for each fluorescence reading during RT-PCR optimization assays and clinical validation. Briefly, the raw fluorescence data was normalized by subtracting the first fluorescence reading. On the basis of the fluorescence ratios obtained, the highest fluorescence ratio was used to identify the best conditions for RT-PCR (one-step or two-step reactions, reverse transcription time, locally produced enzyme concentrations, sample volume), and heatmaps were plotted over time in order to estimate the best conditions for the CRISPR-Cas12a assay (crRNA, Cas12a and MgCl_2_ concentrations).

Clinical performance parameters such as sensitivity and specificity were calculated based on cutoff values determined by a Receiver Operating Characteristic (ROC) curve analysis (Hanley and McNeil, 1982). Cutoff values with the highest percentage of correctly classified samples were selected for fluorescence ratios and initial velocity for N and CII targets, independently. The estimated sensitivity and specificity, and the area under the ROC curve with respective 95% confidence intervals (CI) were reported for all samples and within each grouping by viral RNA load levels.

#### Supplemental Information

**Supplemental Figure 1:**
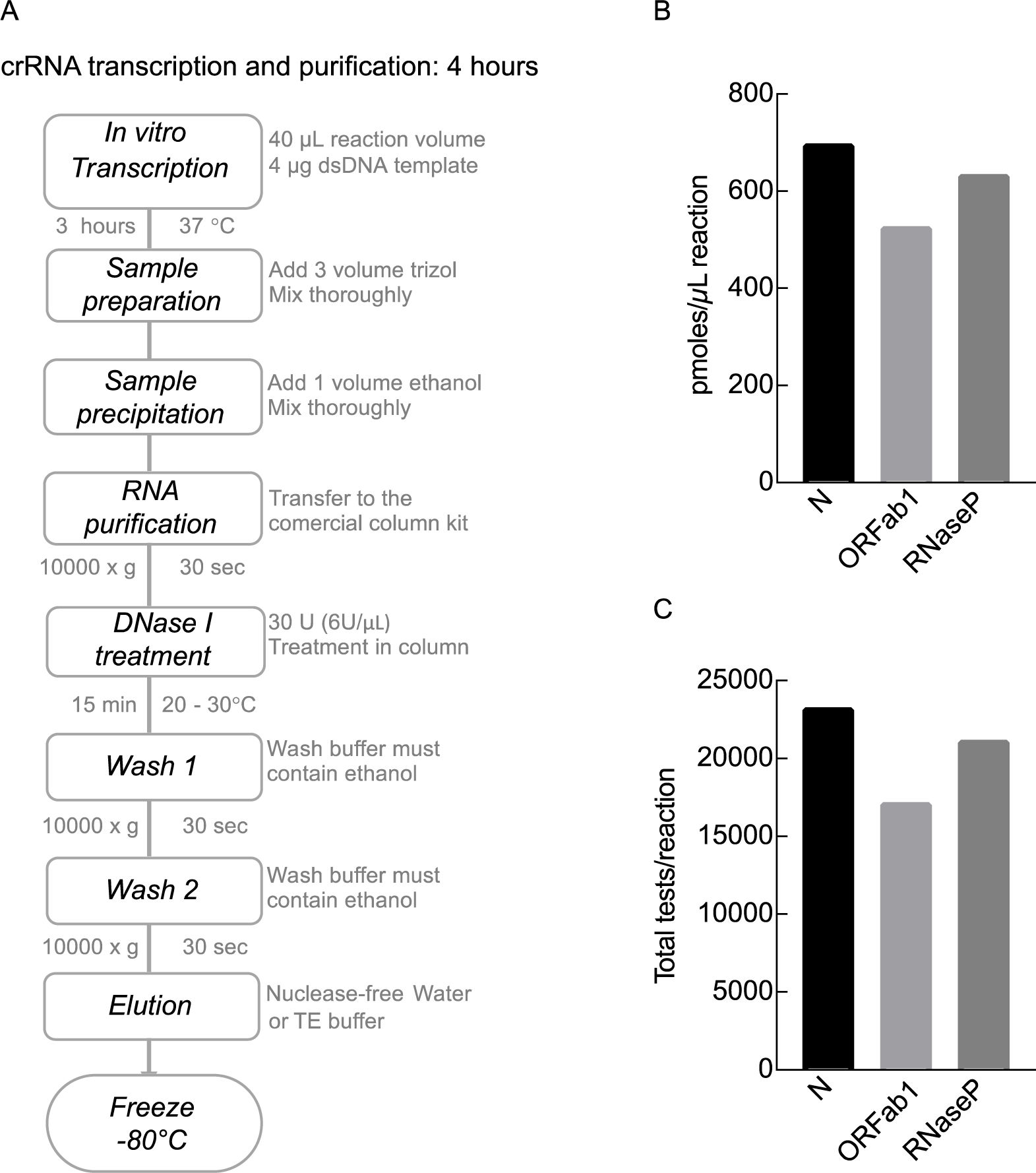
Scheme and results of crRNA production. (A) Production scheme of crRNA from a synthetic dsDNA template using the TranscriptAid T7 High Yield Transcription Kit (Thermo Fisher). (B) Comparison of crRNA yield as the number of crRNA picomoles per microliter of *in vitro* transcription reaction. (C) Estimated number of total CRISPR-Cas12a reactions achieved per each *in vitro* transcription batch of crRNA.

**Supplemental Figure 2:**
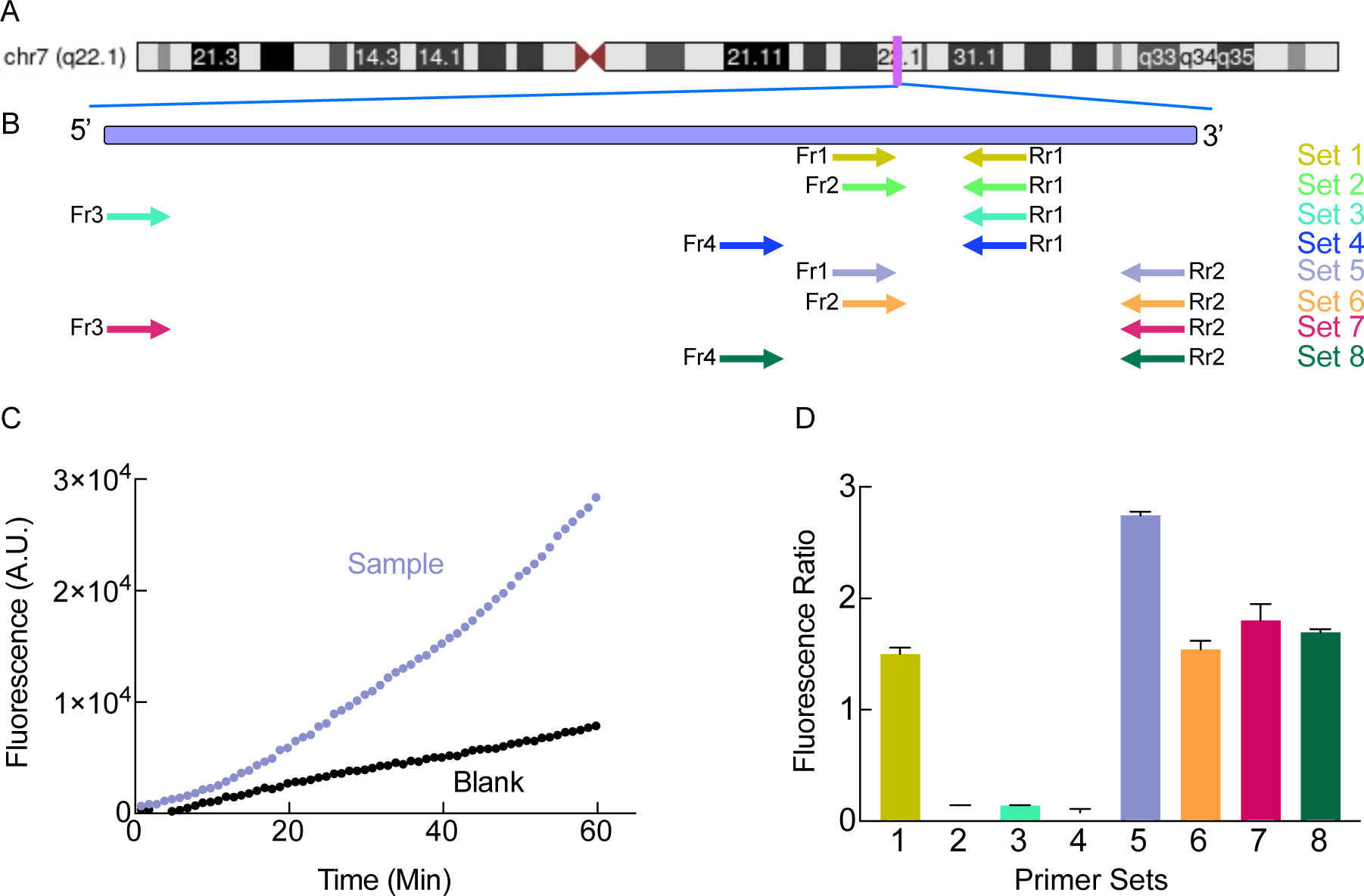
Localization of RNaseP primer sets and RT-PCR efficiency comparison by CRISPR/Cas. (A) Schematic representation of human chromosome 7. The *POP7* gene encoding RNase P protein subunit p20 is located at position 7q22.1. (B) Schematic diagram showing the localization of primer sets for human RNase P *POP7* gene amplification. (C) Fluorescence signal as a function of time for the best primer set (Set 5). A.U., arbitrary units. The RT-PCR non-template control is shown in black symbols (Blank).(D) Comparison of the fluorescence ratio for all eight primer sets. The average fluorescence ratio at 30 minutes is plotted. Error bars show SD of the mean of five measurements.

**Supplemental Figure 3:**
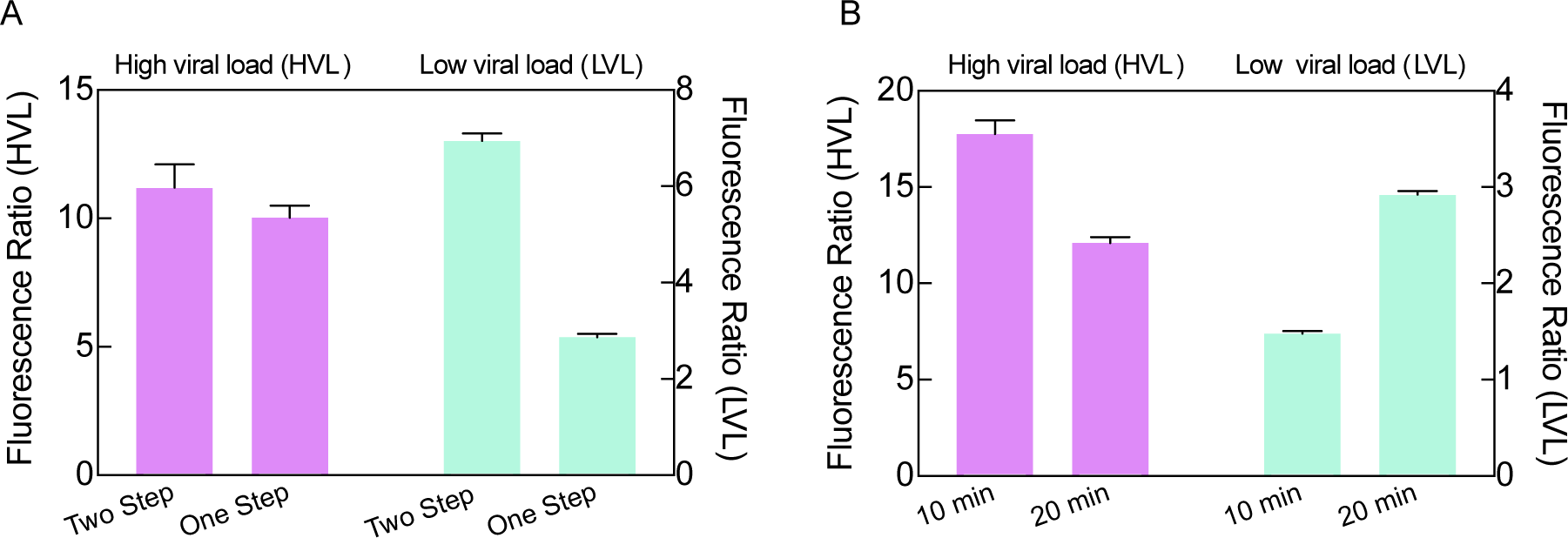
RT-PCR performance comparison by CRISPR/Cas fluorescence detection. (A) One-Step and Two-Step RT-PCR comparison for pooled samples with high or low viral RNA load levels. The average fluorescence ratio at 30 minutes is presented. (B) Same as panel (A) but comparing reverse transcription extension time in One-Step RT-PCR. Error bars show SD of the mean of five measurements.

**Supplemental Figure 4:**
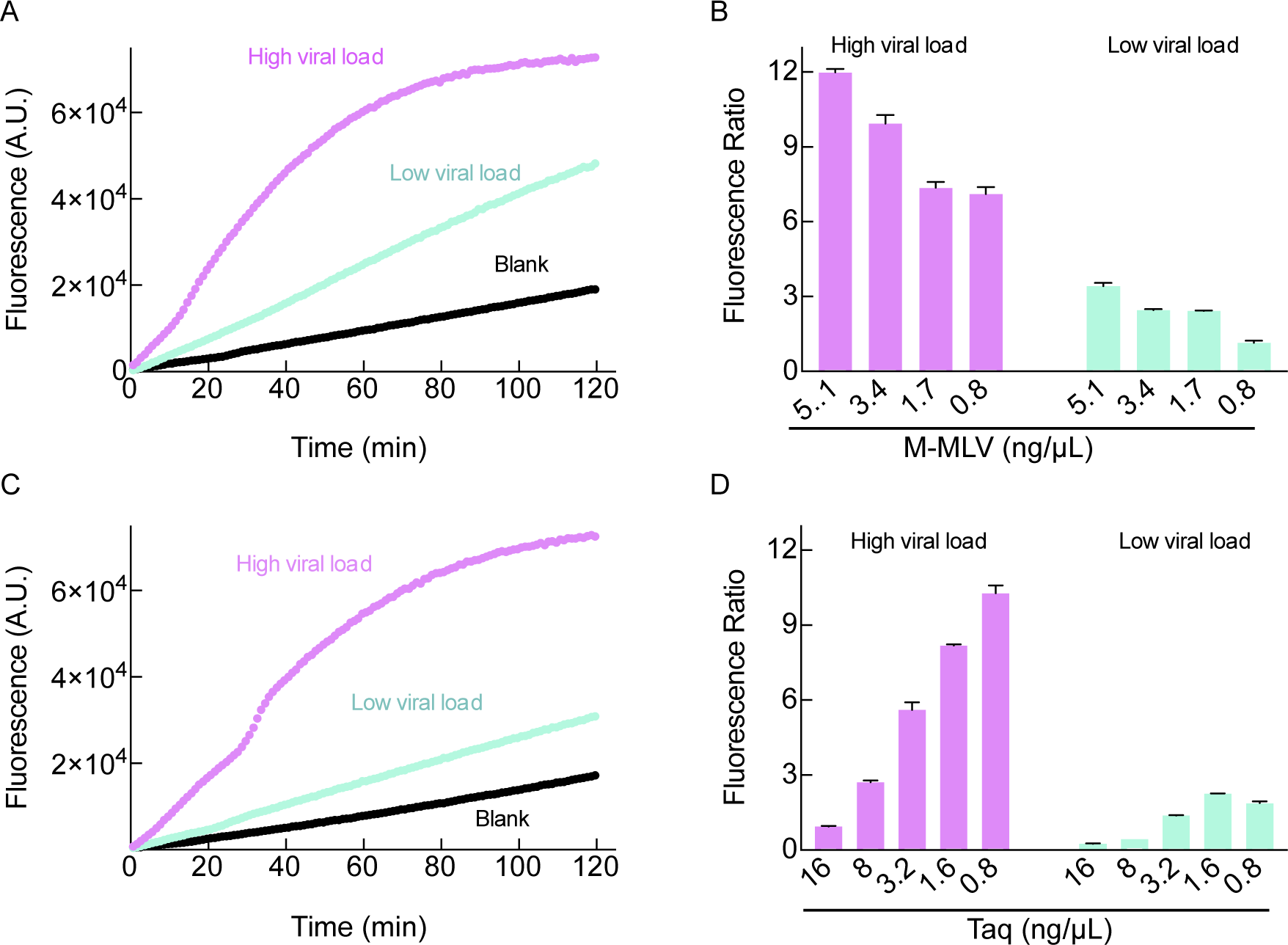
Titrations of M-MLV reverse transcriptase and Taq DNA polymerase. (A) Fluorescence signal (Y-axis) as a function of time for the selected M-MLV reverse transcriptase working concentration (1.7 ng/µL). Samples with high viral load (pink) and low viral load (aquamarine) are compared to the RT-PCR non-template control (black trace, blank). (B) Comparison of the fluorescence ratio (target reaction fluorescence of the test sample to that of the blank reaction) as a function of M-MLV concentration (ng/µL). (C) Same as panel (A) but for the Taq DNA polymerase working concentration (1.6 ng/µL). (D) Same as panel (B) a function of Taq DNA polymerase concentration (ng/µL). All conditions were evaluated using pooled samples with high or low viral RNA load in One-Step RT-PCR. Error bars in (B) and (D) show SD of the mean of five measurements.

**Supplemental Figure 5:**
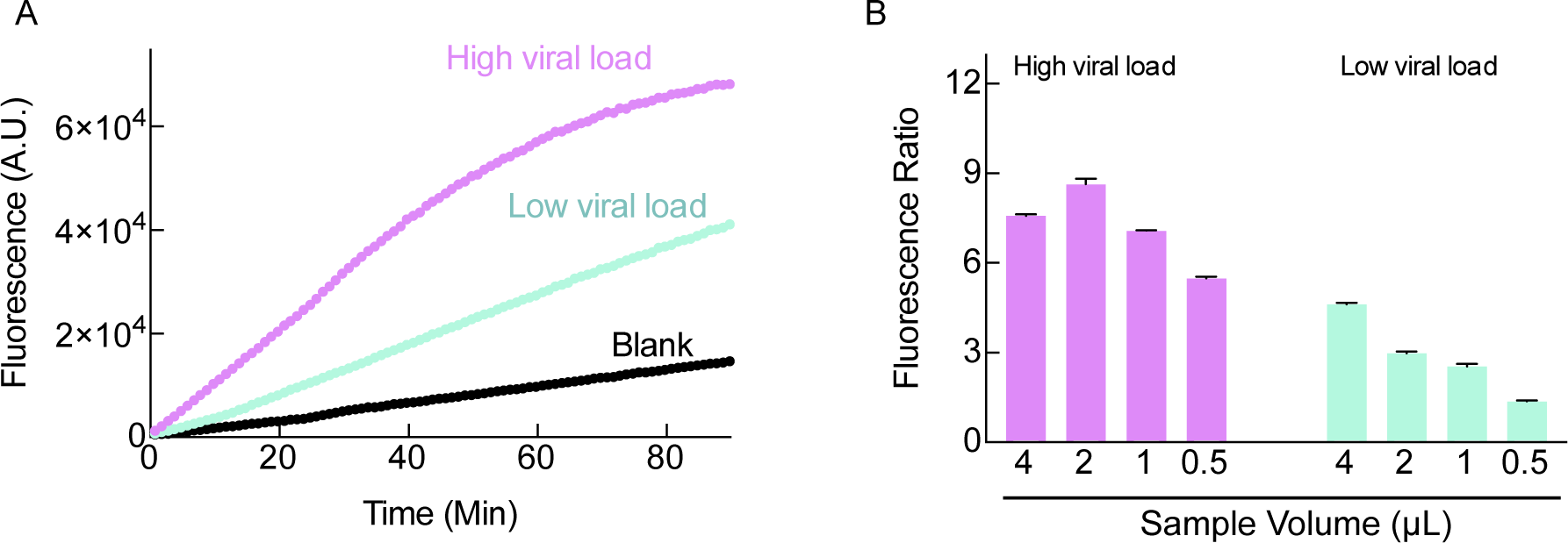
Evaluation of sample volume in the One-Step RT-PCR setup. (A) Fluorescence signal as a function of time using a sample volume of 2 µL in a 20 µL RT-PCR reaction. (B) Comparison of the fluorescence ratio for different sample volumes. Pooled samples with high or low viral RNA load were analyzed. Error bars show SD of the mean of five measurements.

**Supplemental Figure 6:**
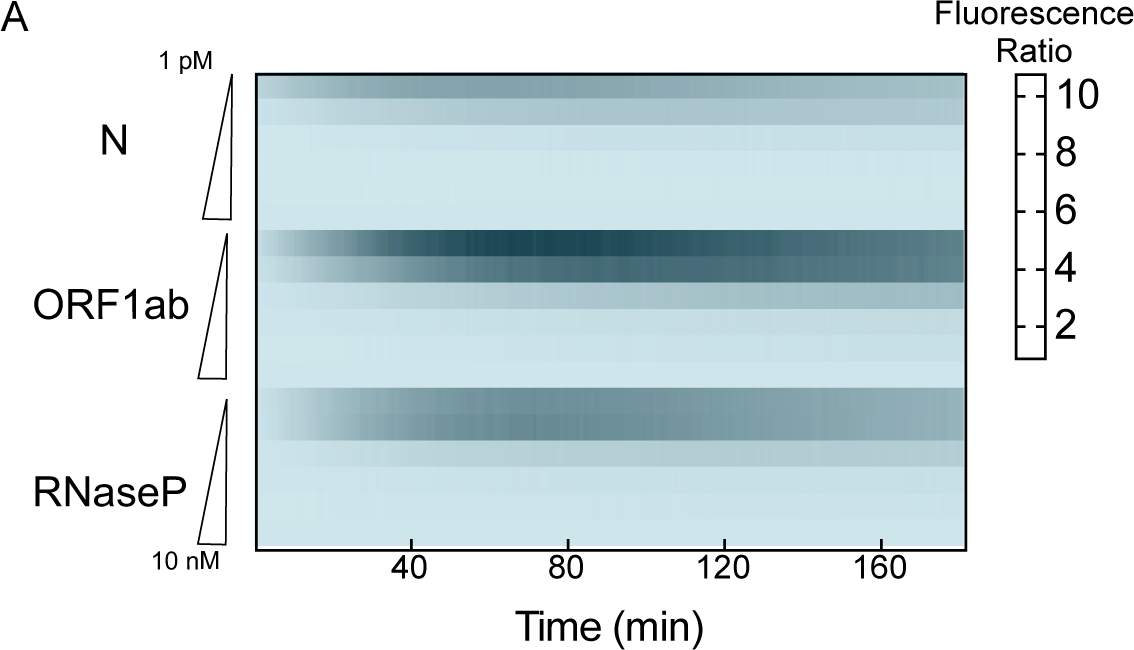
Heatmap of fluorescence ratios dependence on template concentration for both SARS-CoV-2 targets, ORF1ab and N targets, and the human RNaseP target used as sample extraction quality control. Cas12a, crRNA and MgCl_2_ concentrations were 10 nM, 15 nM, and 10 mM, respectively.

**Supplemental Figure 7:**
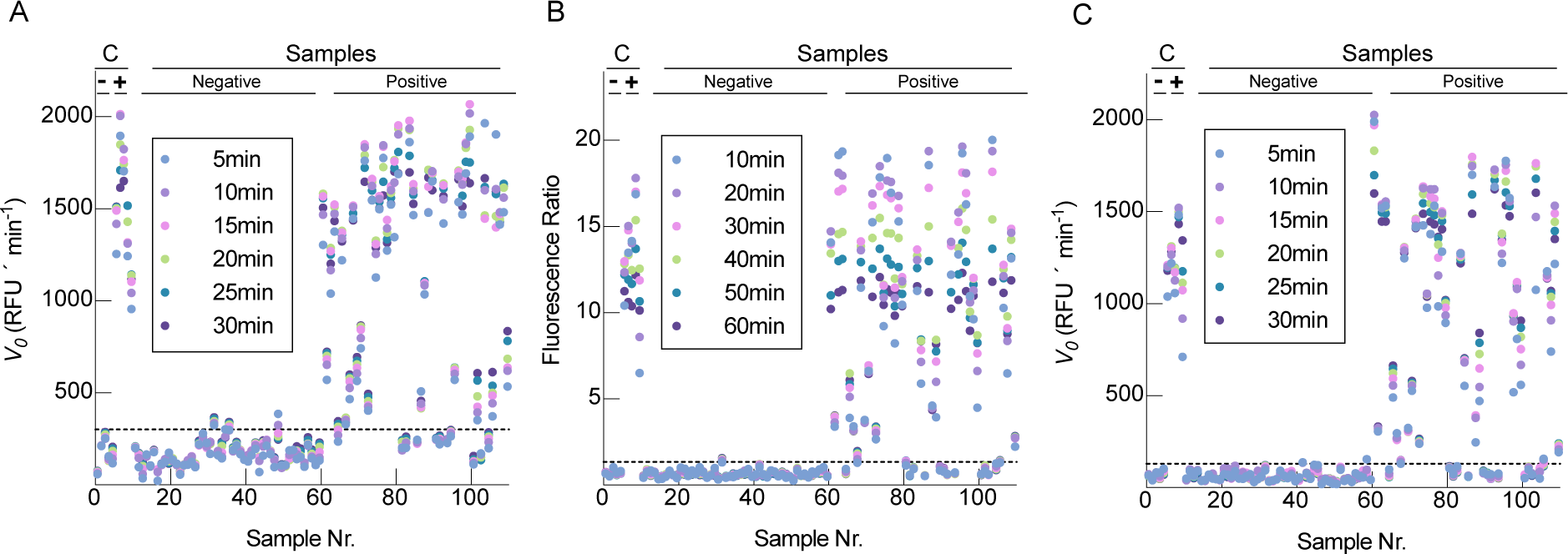
Scatter plot of fluorescence ratios and initial velocity (V_0_) analysis. (A) Distribution of initial velocity values for the N target. (B) and (C) Distribution of fluorescence ratios and initial velocity values, respectively, for the ORF1ab target. Data for clinical samples and controls that tested positive or negative are presented for six time points. Dash lines indicate the thresholds as calculated by the ROC analysis (see results and methods)

**Supplemental Figure 8:**
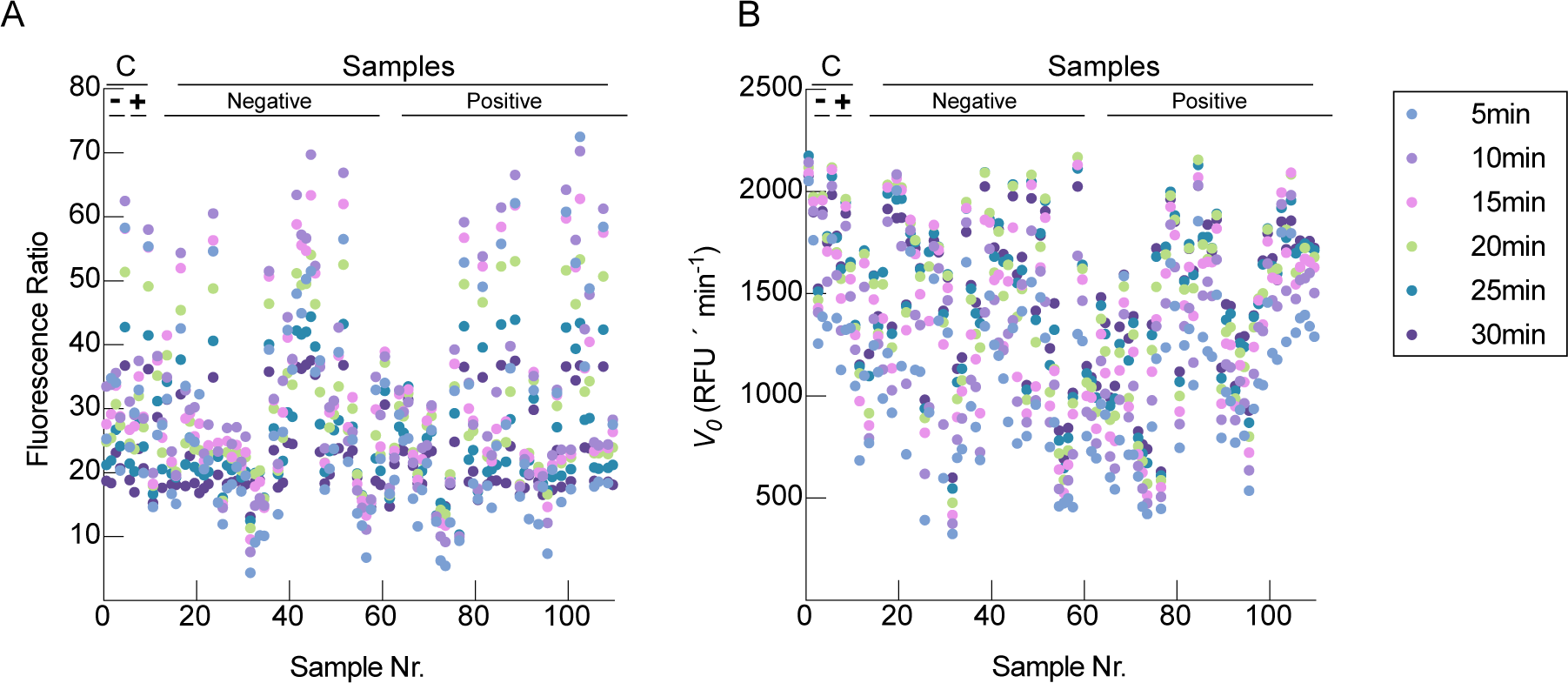
Scatter plot of the fluorescence ratios and initial velocity (V_0_) of the human RNaseP. (A) Distribution of fluorescence ratios. (B) Same as panel (A) but for initial velocity. Data for clinical samples and controls are presented for six time points.

**Supplemental Figure 9:**
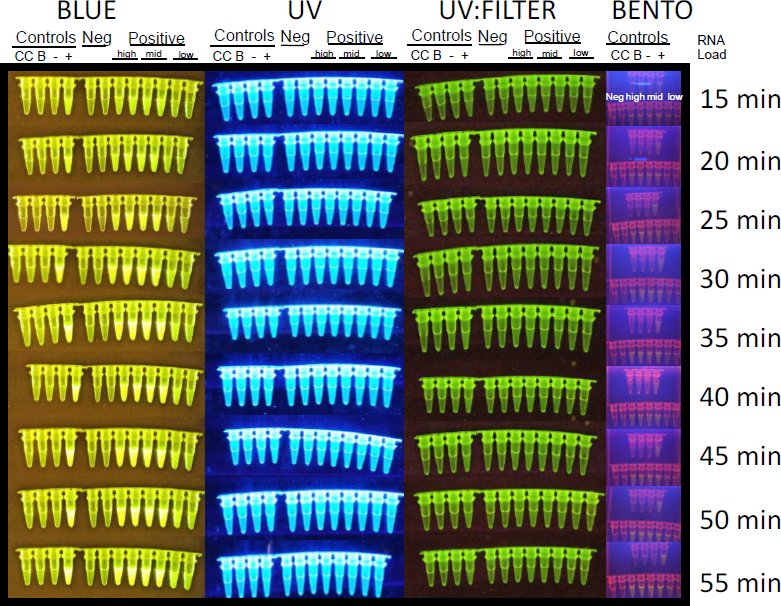
Direct visualization of fluorescence using different transilluminators and the portable molecular equipment BentoLab. Readouts were recorded every five minutes with a blue light transilluminator (270 nm), UV transilluminator without and with filter, and the BentoLab transilluminator. Two clinical samples each at three ranges of viral RNA load levels (high, medium and low), and two negative samples were selected for the assay using the N target. Controls included the CRISPR/Cas12a background control (CC), the non-template RT-PCR control (blank, B), a pool of 10 negative (SARS-CoV-2 not detected) clinical samples, and a pool of 10 positive (SARS-CoV-2 detected with RT-qPCR) clinical samples.

**Supplemental Figure 10:**
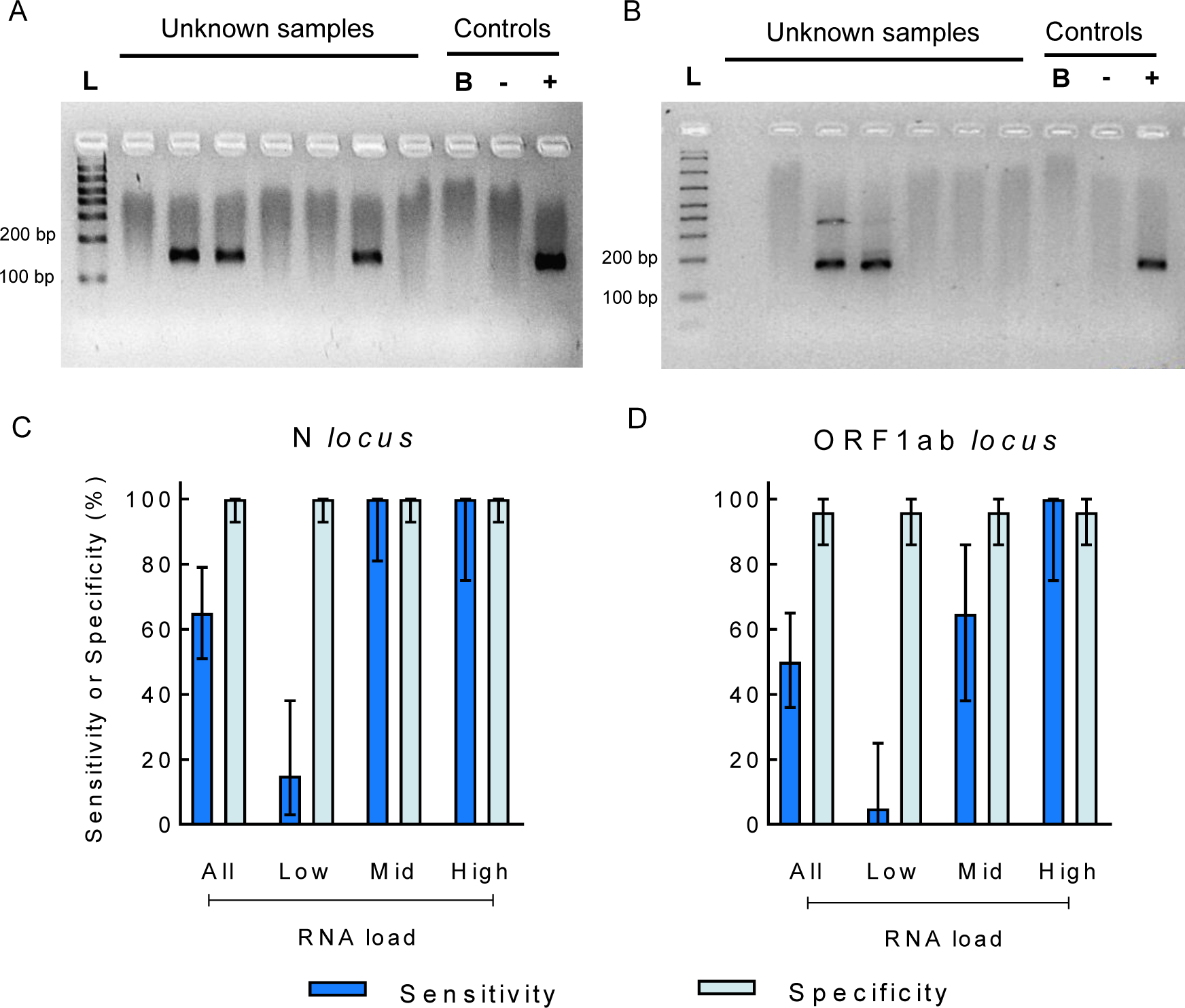
Visualization of RT-PCR products in agarose gel (5%) and diagnostic performance as evaluated by electrophoresis. (A) Amplification products for the N target for seven unknown clinical samples. (B) Same as panel (A) for the ORF1ab target. A blank reaction and pooled positive and negative samples were used as controls. (C) Sensitivity and specificity values with 95% CI estimated for gel visualization readouts for the N target. (D) Same as panel (C) for the ORF1ab target. The sensitivity and specificity data are shown for the complete sample set and within each grouping by viral RNA load levels.

**Supplemental Table 1.** Diagnostic performance as compared to the RT-qPCR gold standard

|  | N Target |  |  | ORF1ab Target |  |  |
| --- | --- | --- | --- | --- | --- | --- |
|  | Sensitivity, %<br>(95% CI) | Specificity, %<br>(95% CI) | ROC area<br>(95% CI) | Sensitivity, %<br>(95% CI) | Specificity, %<br>(95% CI) | ROC area<br>(95% CI) |
| Fluorescence<br>Ratio analysis | 80 (66 – 90) | 100 (93 – 100) | 0.95 (0.91 – 0.99) | 84 (71 – 93) | 96 (86 – 100) | 0.93 (0.87 – 0.98) |
| V<sub>0</sub> analysis | 88 (76 – 96) | 96 (86 – 100) | 0.96 (0.92 – 0.99) | 86 (73 – 94) | 90 (78 – 97) | 0.93 (0.88 – 0.98) |
| Electrophoresis | 66 (51 – 79) | 100 (93 – 100) | 0.83 (0.76 – 0.90) | 50 (36 – 65) | 96 (86 – 100) | 0.73 (0.66 – 0.81) |

Cutoff values were calculated by ROC curve analysis. The estimated sensitivity and specificity, and the area under the ROC curve with 95% confidence intervals (95% CI) are shown for SARS-CoV-2 N and ORF1ab targets.

**Supplemental Table 2.** Diagnostic performance as a function of viral RNA load

|  | N Target |  |  | ORF1ab Target |  |  |
| --- | --- | --- | --- | --- | --- | --- |
|  | Sensitivity, %<br>(95% CI) | Specificity, %<br>(95% CI) | ROC area<br>(95% CI) | Sensitivity, %<br>(95% CI) | Specificity, %<br>(95% CI) | ROC area<br>(95% CI) |
| Low viral load (n = 20) |  |  |  |  |  |  |
| Ratio analysis | 50 (27 – 73) | 100 (93 – 100) | 0.75 (0.64 – 0.86) | 65 (41 – 85) | 96 (86 – 100) | 0.81 (0.70 – 0.92) |
| V<sub>0</sub> analysis | 70 (46 – 88) | 96 (86 – 100) | 0.83 (0.72 – 0.94) | 65 (41 – 85) | 90 (78 – 97) | 0.78 (0.66 – 0.89) |
| Electrophoresis | 15 (3 – 38) | 100 (93 – 100) | 0.58 (0.50 – 0.66) | 5 (0.13 – 25) | 96 (86 – 100) | 0.51 (0.45 – 0.56) |
| Medium viral load (n = 17) |  |  |  |  |  |  |
| Ratio analysis | 100 (81 – 100) | 100 (93 – 100) | 1 | 100 (81 – 100) | 96 (86 – 100) | 0.98 (0.95 – 1) |
| V<sub>0</sub> analysis | 100 (81 – 100) | 96 (86 – 100) | 0.98 (0.95 – 1) | 100 (81 – 100) | 90 (78 – 97) | 0.95 (0.91 – 0.99) |
| Electrophoresis | 100 (81 – 100) | 100 (93 – 100) | 1 | 64.70 (38 – 86) | 96 (86 – 100) | 0.80 (0.68 – 0.92) |
| High viral load (n = 13) |  |  |  |  |  |  |
| Ratio analysis | 100 (75 – 100) | 100 (93 – 100) | 1 | 92 (64 – 100) | 96 (86 – 100) | 0.94 (0.86 – 1) |
| V<sub>0</sub> analysis | 100 (75 – 100) | 96 (86 – 100) | 0.98 (0.95 – 1) | 100 (75 – 100) | 90 (78 – 97) | 0.95 (0.91 – 0.99) |
| Electrophoresis | 100 (75 – 100) | 100 (93 – 100) | 1 | 100 (75 – 100) | 96 (86 – 100) | 0.98 (0.95 – 1) |

Cutoff values were calculated by ROC curve analysis. The estimated sensitivity and specificity, and the area under the ROC curve with 95% confidence intervals (95% CI) are shown for SARS-CoV-2 N and ORF1ab targets for three viral RNA load levels.

**Supplemental Table 3.** Comparison of the test performance with two methods for cut-off calculation

|  |  | N Target |  |  | ORF1ab Target |  |  |
| --- | --- | --- | --- | --- | --- | --- | --- |
|  |  | Sensitivity, %<br>(95% CI) | Specificity, %<br>(95% CI) | ROC area<br>(95% CI) | Sensitivity, %<br>(95% CI) | Specificity, %<br>(95% CI) | ROC area<br>(95% CI) |
| Fluorescence<br>ratio analysis | ROC analysis | 80 (66 – 90) | 100 (93 – 100) | 0.95 (0.91 – 0.99) | 84 (71 – 93) | 96 (86 – 100) | 0.93 (0.87 – 0.98) |
|  | mean + 3 SD | 78 (64 – 89) | 100 (93 – 100) | 0.89 (0.83 – 0.95) | 76 (62 – 87) | 96 (86 – 100) | 0.86 (0.79 – 0.93) |
| Initial velocity<br>analysis | ROC analysis | 88 (76 – 96) | 96 (86 – 100) | 0.96 (0.92 – 0.99) | 86 (73 – 94) | 90 (78 – 97) | 0.93 (0.88 – 0.98) |
|  | mean + 3 SD | 76 (62 – 87) | 100 (93 -100) | 0.88 (0.82 – 0.94) | 78 (64 – 89) | 94 (84 – 99) | 0.86 (0.79 – 0.93) |

Fluorescence ratio and initial velocity data of the CRISPR-Cas12a assays were analyzed and the cutoff values were calculated by ROC curve or taken as the mean of 10 negative controls plus 3 times the standard deviation (SD). The estimated sensitivity and specificity, and the area under the ROC curve with 95% confidence intervals (95%CI) are shown for SARS-CoV-2 N and ORF1ab targets.

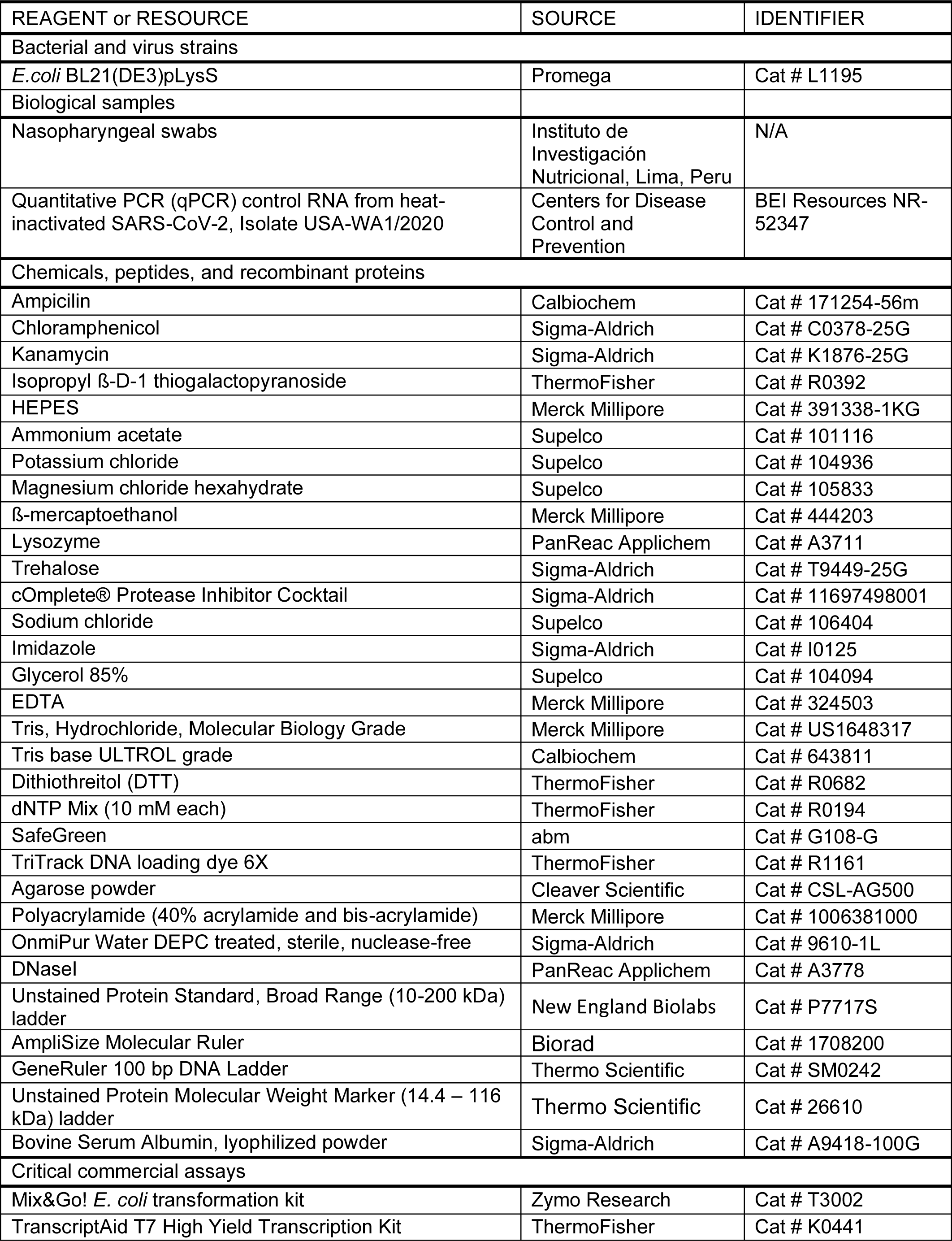
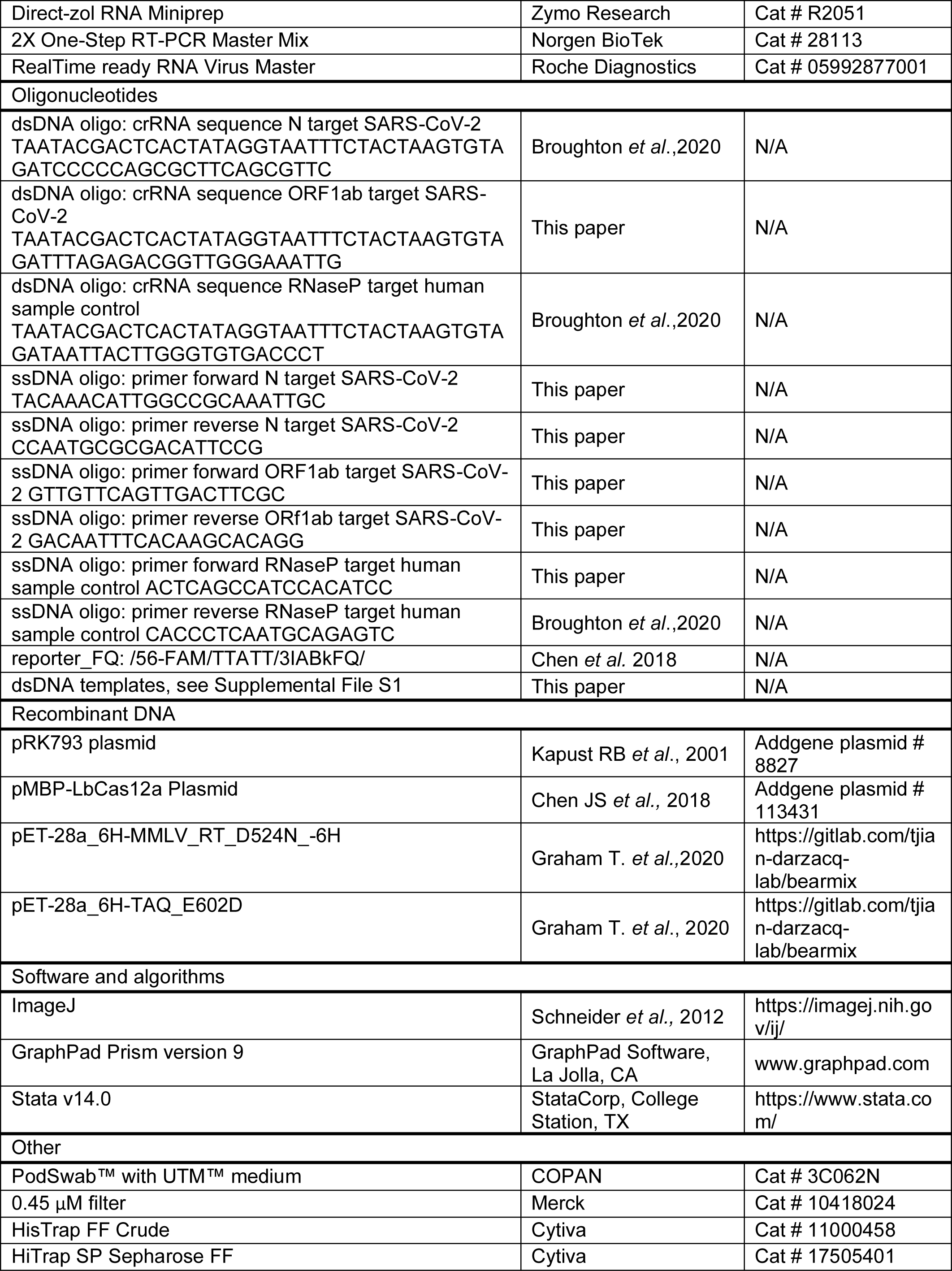
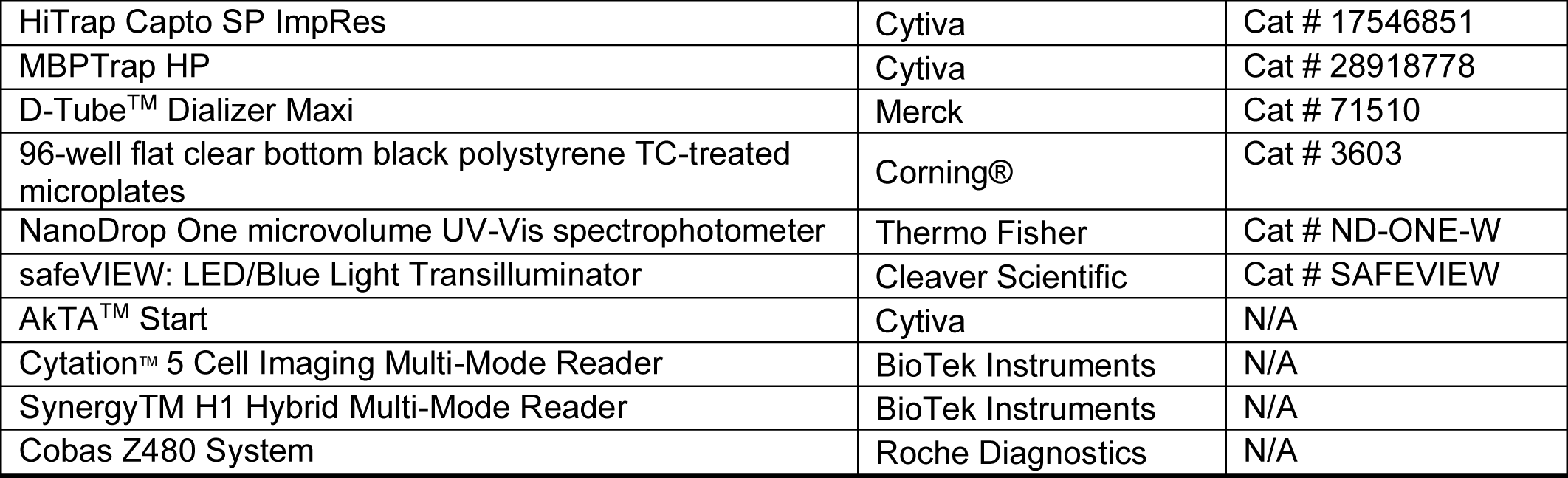
KEY RESOURCES TABLE

Supplemental File S1: Excel sheet with construct sequences, crRNAs, and primers

Supplemental File S2: Alignment file of SARS-CoV-2 genomes

